# Genome-wide association study of chronic sputum production implicates loci involved in mucus production and infection

**DOI:** 10.1101/2022.01.11.22269075

**Authors:** RJ Packer, N Shrine, R Hall, CA Melbourne, R Thompson, AT Williams, ML Paynton, AL Guyatt, PH Lee, C John, A Campbell, C Hayward, M de Vries, JM Vonk, J Davitte, E Hessel, D Michalovich, JC Betts, I Sayers, A Yeo, IP Hall, MD Tobin, LV Wain

**Affiliations:** Department of Health Sciences, University of Leicester, Leicester, UK; Centre for Respiratory Research, NIHR Nottingham Biomedical Research Centre, School of Medicine, Biodiscovery Institute, University of Nottingham, Nottingham, UK; Centre for Genomic and Experimental Medicine, Institute of Genetics & Cancer, University of Edinburgh, Western General Hospital, Edinburgh EH4 2XU, UK; Medical Research Council Human Genetics Unit, Institute of Genetics and Cancer, University of Edinburgh, Edinburgh EH4 2XU, UK; University of Groningen, University Medical Center Groningen, Department of Epidemiology & Groningen Research Institute for Asthma and COPD (GRIAC), Groningen, The Netherlands; GSK R&D, Collegeville, PA, USA; GSK R&D, Stevenage, UK; Leicester NIHR Biomedical Research Centre, Glenfield Hospital, Leicester, UK

## Abstract

**Background:** Chronic sputum production impacts on quality of life and is a feature of many respiratory diseases. Identification of the genetic variants associated with chronic sputum production in a disease agnostic sample could improve understanding of its causes and identify new molecular targets for treatment.

**Methods:** We conducted a genome-wide association study (GWAS) of chronic sputum production in UK Biobank. Signals meeting genome-wide significance (P<5×10^−8^) were investigated in additional independent studies, were fine-mapped, and putative causal genes identified by gene expression analysis. GWAS of respiratory traits were interrogated to identify whether the signals were driven by existing respiratory disease amongst the cases and variants were further investigated for wider pleiotropic effects using phenome-wide association studies (PheWAS).

**Findings:** From a GWAS of 9,714 cases and 48,471 controls, we identified six novel genome-wide significant signals for chronic sputum production including signals in the Human Leukocyte Antigen (HLA) locus, chromosome 11 mucin locus (containing *MUC2, MUC5AC* and *MUC5B*) and the *FUT2* locus. The four common variant associations were supported by independent studies with a combined sample size of up to 2,203 cases and 17,627 controls. The mucin locus signal had previously been reported for association with moderate-to-severe asthma. The HLA signal was fine-mapped to an amino-acid change of threonine to arginine (frequency 36.8%) in HLA-DRB1 (HLA-*DRB1*\*03:147). The signal near *FUT2* was associated with expression of several genes including *FUT2*, for which the direction of effect was tissue dependent. Our PheWAS identified a wide range of associations.

**Interpretation:** Novel signals at the *FUT2* and mucin loci highlight mucin fucosylation as a driver of chronic sputum production even in the absence of diagnosed respiratory disease and provide genetic support for this pathway as a target for therapeutic intervention.

## Introduction

Increased sputum production impacts on daily activities and quality of life - and is a shared feature of many respiratory diseases. Worldwide, 545 million people have chronic respiratory conditions, with those associated with chronic sputum production including chronic obstructive pulmonary disease (COPD), asthma, bronchiectasis, chronic bronchitis, and cystic fibrosis. Chronic respiratory disease is the third leading cause of death worldwide, with 3.91 million deaths in 2017 [1].

The determinants of chronic sputum production in disease are not completely understood [2]. Most studies of excess sputum production have been in subjects with chronic bronchitis and COPD where it has been associated with lower lung function [3, 4] and higher risk of both exacerbation and respiratory symptoms [5]. Risk factors for excess sputum production include smoking and occupational and environmental pollutants [4, 6–8]. Currently available drug treatments for those with chronic sputum production do not generally affect the rate of production of sputum, but act as mucolytics and expectorants [9–11].

Genome-wide association studies have highlighted pathways underlying a range of respiratory traits and diseases, and highlighted potentially relevant drug targets [12, 13]. Previous genome wide association studies of sputum production [14–17] and have not identified any genome-wide significant findings.

We hypothesised that identifying genetic variants that are associated with chronic sputum production in a large general population sample could improve understanding of its causes and identify new molecular targets for treatment. To test this hypothesis, we undertook a genome-wide association study (GWAS) of risk of chronic sputum production in 9,714 cases and 48,471 controls from UK Biobank and sought replication of the association signals in five additional independent studies totalling 2,203 cases and 17,627 controls. We performed phenome-wide association studies (PheWAS) and interrogation of gene expression data to characterise the association signals and determine which genes may be driving these signals.

## Methods

### Study population

Information about chronic sputum production was obtained from the online lifetime occupation survey that was emailed to 324,653 UK Biobank participants with existing email addresses between June and September 2015 and achieved a response rate of 38% (31% of all of those contacted provided a full completion of the questionnaire [18]). For this study, we defined cases as those who answered “yes” to the question “do you bring up phlegm/sputum/mucus daily?” (UK Biobank data-field 22504, total 121,283 participants provided a “yes” or “no” response). Controls were defined as those who answered “no” to this question. Cases and controls were further restricted to those of genetically-determined European ancestry, as previously defined [19], with available smoking data (data-field 20160). Related individuals were removed, with cases preserved over controls when excluding one of a pair (or more) of related individuals (data-field 22021, “related” defined as a KING kinship coefficient ≥ 0.0884, equivalent to second-degree relatedness or closer). For related pairs within the cases or controls, the individual with the lowest genotype missingness (data-field 22005) was retained. From all available controls, we defined a subset of controls with a similar age (data-field id 34) and sex (data-field id 31) distribution to the cases at a 1:5 ratio with the cases.

Demographics and respiratory characteristics of the case and controls were derived using the following definitions: doctor-diagnosed asthma (UK Biobank data-field 22127), moderate-to-severe asthma (as previously described [20]), doctor-diagnosed chronic bronchitis (data-field 22129), cough on most days (data-field 22502), smoking status (data-field 20160), COPD Global Initiative for Chronic Obstructive Lung Disease (GOLD) stage 1-4 and stages 2-4 (defined using baseline spirometry as previously described [19], [21]) bronchiectasis and cystic fibrosis (Supplementary Tables 1 and 2).

UK Biobank has ethical approval from North West – Haydock Research Ethics Committee (21/NW/0157).

### Genome-wide association study of chronic sputum production

Genetic data from the v3 March 2018 UK Biobank data release, imputed to the Haplotype Reference Consortium panel r1.1 2016, was used for the genome-wide association study.

Association testing was performed using logistic regression under an additive genetic model in PLINK 2.0 [22] with age, sex, array version, never/ever smoking status and the first 10 principal components of ancestry as covariates. Variants were excluded if they had an imputation quality INFO score <0.5 or a minor allele count (MAC) <20. Association signals were considered genome-wide significant at P<5×10^−8^. Independent signals were initially defined using a 1Mb window (500kb each side of the sentinel variant) and then using conditional analyses implemented in GCTA-COJO [23]. All variant coordinates are for genome build GRCh37. Region plots were created using LocusZoom [24].

### Replication

We sought replication in five general population cohorts which surveyed participants for chronic sputum production; Generation Scotland [25], EXCEED Study [26], LifeLines 1, LifeLines 2 and Vlagtwedde-Vlaardingen[17]. Further details are provided in the Supplementary text.

In addition, the overlap of primary care sputum codes with the chronic sputum production question (UK Biobank data-field 22504) was evaluated to identify whether primary care codes could be used to define an additional independent case-control dataset from those in UK Biobank who did not respond to the online lifetime occupation survey (Supplementary text).

### Fine-mapping

We undertook Bayesian fine-mapping (29) for all genome-wide significant signals that were not in the HLA region to define 99% credible sets of variants i.e. sets of variants that are 99% probable to contain the true causal variant (assuming that it has been measured).

To fine-map signals within the HLA region (chr6:29,607,078-33,267,103 (b37)) to a specific HLA gene allele or amino acid change, we re-imputed our discovery samples using IMPUTE2 v2.3.1 with a reference panel that enabled imputation of 424 classical HLA alleles and 1,276 amino acid changes as described in [27]. We then repeated the association testing as described above.

### Mapping association signals to putative causal genes

We used functional annotation and co-localisation with expression Quantitative Trait Loci (eQTL) signals to identify putative causal genes at each signal.

Annotation of the variants in each credible set was performed using SIFT [28], PolyPhen-2 and CADD, all implemented using the Ensemble GRCh37 Variant Effect Predictor (VEP) [29], alongside FATHMM [30]. Variants were annotated as deleterious if they were labelled deleterious by SIFT, probably damaging or possibly damaging by PolyPhen-2, damaging by FATHMM (specifying the “Inherited Disease” option of the “Coding Variants” method, and using the “Unweighted” prediction algorithm) or had a CADD scaled score ≥20.

We queried the sentinel variants in GTEx V8 [31] and BLUEPRINT [32] (see Supplementary Table 3 for list of tissues). We tested for colocalisation of GWAS and eQTL signals using coloc [33]; H4 >80% was used to define a shared causal variant for eQTL and GWAS signals.

### Associations with other phenotypes

To investigate whether the signals of association with sputum production were driven by underlying respiratory phenotypes of the cases, a look-up for each signal was undertaken for fourteen respiratory or respiratory-related traits from GWAS results (moderate-to-severe asthma (N cases=5,135, controls=25,675) [20], lung function (Forced Expired Volume in 1 second [FEV_1_], Forced Vital Capacity [FVC], FEV_1_/FVC, peak expiratory flow (PEF)) (N=400,102) [19], respiratory infection (N cases=19,459, controls=101,438) [34], chronic cough (N cases=15,213, controls=94,731), chronic bronchitis (N cases=977, controls = 108,967), idiopathic pulmonary fibrosis (IPF) (N cases=2,668, controls=8,591) [35], smoking traits (Smoking age-of-onset (N=124,590), smoking cessation (N cases=141,649, controls=27321), smoking cigarettes-per-day (N=120,744), smoking initiation (N cases=170,772, controls=212,859) and asthma (N cases=23,948, controls=118,538) [36]). Smoking trait results were from the UK Biobank component of [37]; chronic cough and chronic bronchitis were defined for this study using UK Biobank data, see Supplementary text. Where the sentinel variant was not available in the look-up dataset, we utilised an alternative variant from the credible set with the highest posterior probability of being causal. A Bonferroni adjustment for 84 association tests was applied requiring a P <5.95×10^−4^ for association to be classified as statistically significant. Imputed HLA gene allele or amino acid changes were used for signals in the HLA region.

To investigate associations of the chronic sputum-associated variants with a wider range of phenotypes, we performed PheWAS for 2,172 traits in UK Biobank (FDR<0.01, Supplementary Text) and searched the Open Targets Genetics Portal (P<5×10^−8^, version 0.4.0 (bd664ca) - accessed 16^th^ April 2021[38]). PheWAS for imputed HLA alleles was performed using DeepPheWAS [39] (see Supplementary text).

### Sensitivity analyses

To further investigate whether the effects of the variants associated with risk of chronic sputum production differ between ever and never smokers, or between individuals with and without a history of chronic respiratory disease (spirometry defined COPD GOLD1+, doctor diagnosed asthma or doctor diagnosed chronic bronchitis), we tested association of sentinel variants in ever and never smokers and those with and without evidence of chronic respiratory disease separately. We additionally evaluated whether the associations differed between males and females or by the time of year of the survey (UK Biobank data-field 22500). Finally, we evaluated whether adjusting for current smoking (UK Biobank data-field 22506) (rather than ever vs never smoker status) affected the results.

## Results

A total of 10,481 participants answered “yes” to the question “Do you bring up phlegm/sputum/mucus daily?” and 110,802 answered “no” (Supplementary Table 4). After excluding those with missing genotype and essential covariate data, and those of genetically-determined European ancestry, a total of 9,714 cases and 48,471 controls (Figure 1), and 27,317,434 variants, were included in the GWAS. Ever-smoking and respiratory disease were more common in the cases than in the controls (Table 1). The genomic control inflation factor (lambda) was 1.026 so no adjustments to the test statistics were applied (Supplementary Figure 1). Six independent novel signals met the genome-wide significance threshold of P<5×10^−8^ (Figure 2 and Table 2). These were four common variant signals (minor allele frequency > 5%) in or near *MUC2, FUT2*, HLA*-DRB1* and *NKX3-1*, and two intronic rare variant signals (minor allele frequency < 1%) in *OCIAD1* and *NELL1* (Supplementary Figures 2 to 7).

**Figure 1.**
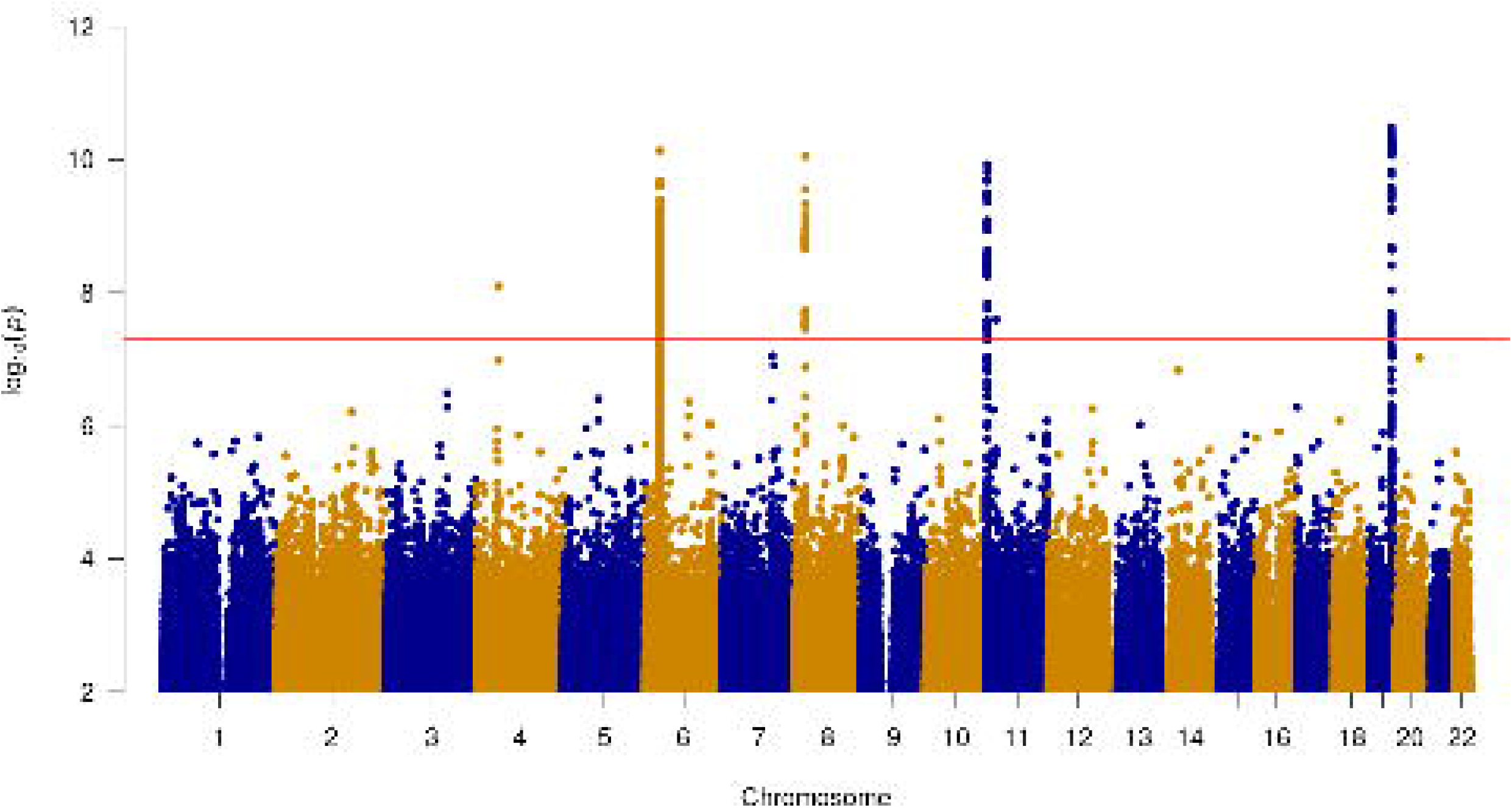
Study flow chart detailing case control selection from the UK Biobank cohort.

**Table 1.** Demographics, ever-smoking status, doctor-diagnosed asthma, doctor-diagnosed chronic bronchitis, cough, moderate-to-severe asthma and COPD GOLD stage 1-4 status of cases and controls included in the GWAS of chronic sputum production. *Total 6942 cases and 36321 controls with available spirometry that passed QC.

|  | Cases<br>N=9714 | Controls<br>N=48471 |
| --- | --- | --- |
| Mean age (years) | 57.7 | 57.7 |
| % female | 42.5 | 42.5 |
| Ever smoked (%) | 5306 **(54.6)** | 20912 **(43.1)** |
| Current smoker (%) | 983 **(10.2)** | 1569**(3.2)** |
| Doctor-diagnosed chronic bronchitis (%) | 407 **(4.2)** | 416 **(0.86)** |
| Doctor-diagnosed asthma (%) | 2630 **(27.1)** | 5251 **(10.8)** |
| Cough on most days (%) | 7022 **(72.3)** | 3999 **(8.3)** |
| Moderate-to-severe asthma (%) | 520 **(5.4)** | 521 **(1.1)** |
| Meets spirometry criteria GOLD 1-4 (%) | 1511 **(21.8)\*** | 4766 **(13.1)\*** |

**Figure 2.**
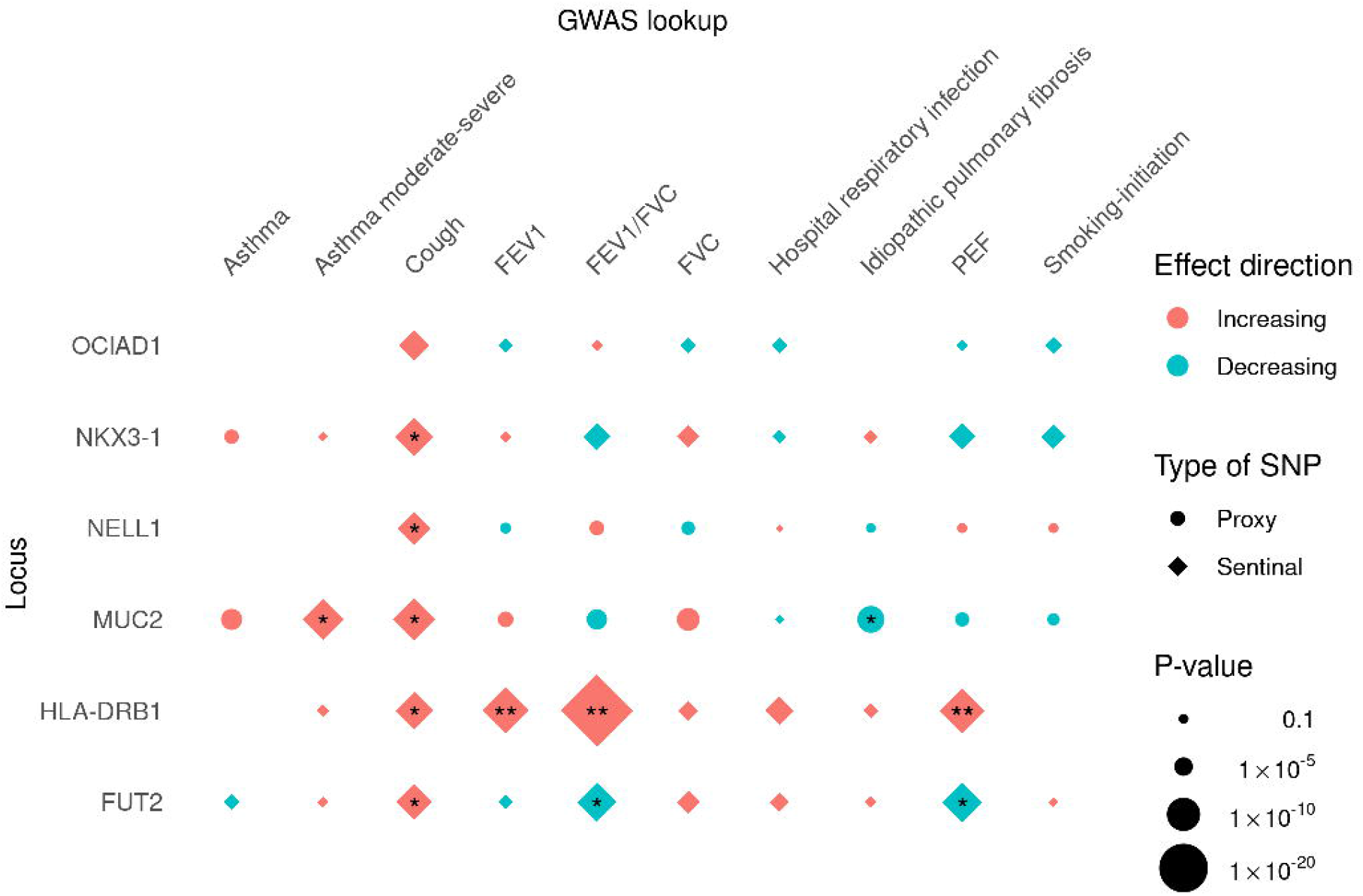
Manhattan plot for the genome-wide association study of chronic sputum production. The red line indicates genome-wide significance.

**Table 2:** Novel genome-wide significant signals of association with chronic sputum production.

| Chr:Position (GRCh37) | RSID | Locus (BP distance from gene)* | coded/non-coded | Coded allele frequency % (count) | OR (95 CI) | P | Imputation quality (INFO) | # variants in 99% credible set (highest posterior probability) |
| --- | --- | --- | --- | --- | --- | --- | --- | --- |
| 4:48854355 | rs79998532 | *OCIAD1* (intronic) | A/G | 0.2% (233) | 2.36 (1.76-3.16) | $8.00 \times 10^{-09}$ | 0.92 | 3 (0.86) |
| 6:32496534 | rs374248993 | *HLA-DRB1* ☐ | G/C | 57% (66355) | 1.12 (1.08-1.16) | $7.30 \times 10^{-11}$ | 0.87 | *HLA-DRB1* *03:147☐ |
| 8:23480686 | rs79401075 | *NKX3-1* (59,765) | A/G | 10% (11620) | 1.18 (1.12-1.24) | $8.90 \times 10^{-11}$ | 0.98 | 30 (0.32) |
| 11:1116931 | rs779167905 | *MUC2* (12,513) | T/TTCTA | 67% (78158) | 1.12 (1.08-1.16) | $1.20 \times 10^{-10}$ | 0.98 | 30 (0.15) |
| 11:20887601 | rs529240826 | *NELL1* (intronic) | GC/G | 0.51% (588) | 1.91 (1.52-2.4) | $2.50 \times 10^{-08}$ | 0.67 | 2 (0.83) |
| 19:49206417 | rs492602 | *FUT2* (exonic) | G/A | 51% (58803) | 1.11 (1.08-1.15) | $3.20 \times 10^{-11}$ | 1 | 32 (0.07) |

☐ = amino-acid change of threonine to arginine at codon 233 of exon 5 of *HLA-DRB1* (HLA gene allele *HLA-DRB1*\*03:147)
BP = base pairs; OR = odds ratio; CI = confidence interval
\*Start or end of nearest gene

No systematic differences were seen in effect sizes when stratifying by smoking status, by history of chronic respiratory disease, by sex, by time of year of survey or when including current smoking status as a covariate (Supplementary Table 5, Supplementary Figures 8 to 13) for the six sentinel variants. Through comparison of survey responses and linked primary care data we showed that primary care codes were not adequate proxies for the survey responses (Supplementary Text). We sought replication in five independent cohorts with a combined sample size to 1977 cases and 17,627 controls; data from all five replication cohorts were only available for the *FUT2* locus. Although none of the signals met criteria for significance in a meta-analysis of the replication cohorts, the directions of effect were consistent with the discovery results for the signals in or near *MUC2, FUT2, OCIAD1*, HLA*-DRB1* and *NKX3-1* and all except the signals at *NELL1* and HLA*-DRB1* also increased in significance when the replication and discovery results were meta-analysed (Supplementary Table 13 and Supplementary Figure 14).

### Novel associations with chronic sputum production

#### HLA *locus*

The HLA signal was fine-mapped to an amino-acid change of threonine to arginine (frequency 36.8%) at codon 233 of exon 5 of *HLA-DRB1* (HLA-*DRB1*\*03:147) that was associated with decreased risk (OR 0.91 [95% C.I. 0.88-0.94]) of chronic sputum production (P=3.43×10^−9^). The amino acid change was in linkage disequilibrium with the GWAS sentinel variant rs374248993 (R^2^=0.74) and the signal for rs374248993 was attenuated when conditioned on the amino acid change (Supplementary Figures 15 and 16).

HLA-*DRB1*\*03:147 was significantly associated with FEV_1_, FEV_1_/FVC and PEF at genome-wide significance (P<5×10^−8^) (Figure 3 and Supplementary Table 6). The amino acid associated with increased risk of chronic sputum production (threonine) was associated with increased lung function; this had not been previously reported. The HLA PheWAS identified multiple significant associations for the HLA allele associated with increased risk of chronic sputum production with a wide range of quantitative traits (for example, blood cell traits, liver biomarkers) and diseases (including decreased risk of gastrointestinal and thyroid-associated diseases, and increased risk of bronchiectasis and asthma) (Supplementary Table 7).

**Figure 3.**
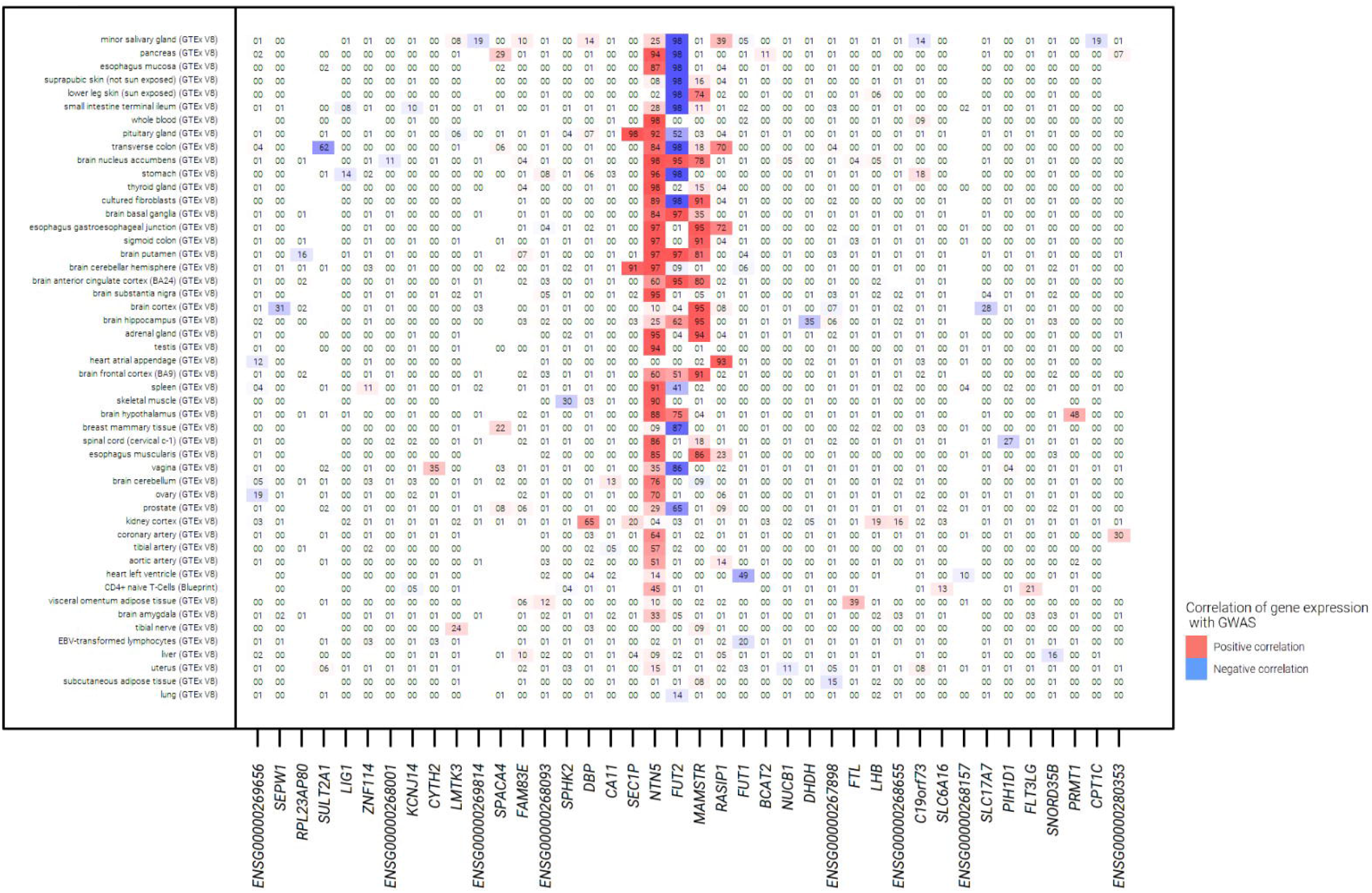
Results for association of sentinel variant risk alleles with respiratory traits. Results are aligned to the risk allele for chronic sputum production, effect direction ‘Increasing’ can be read as increasing risk for binary traits and increasing values in quantitative traits. Chronic bronchitis and smoking age of onset, cigarettes per day and cessation phenotype lookups were omitted as no associations with P<0.05 found. *P <5.95×10^−4^ (Bonferroni adjustment for 84 association tests) ** P<5×10^−8^.

#### MUC2 locus

For the mucin locus signal (rs779167905 allele), the allele associated with risk of chronic sputum production was also significantly associated with increased risk of asthma (OR 1.06, P=0.0027) and moderate-to-severe asthma (OR 1.13, P=6.3×10^−7^), increased FVC (beta 0.0087, P=6×10^−4^) and decreased risk of IPF (OR 0.84, P=7.5×10^−6^) (Figure 3, Supplementary Table 6). There were no associations with gene expression for rs779167905.

Genome-wide significant associations with IPF [40] and moderate-severe asthma [20] have previously been reported at this chromosome 11 locus and so we undertook a conditional analysis to identify whether the chronic sputum production signal was independent of these previous signals. Repeating the association testing for this variant conditioning on the previously reported variants (rs35705950 [40] and rs11603634 [20]) identified that the chronic sputum production GWAS signal was independent of the IPF signal (rs779167905, conditional P=1.18×10^−10^) but was not independent of the previously reported moderate-to-severe asthma signal (rs779167905, conditional P=0.0039) (Supplementary Figures 17 and 18).

Our PheWAS and Open Targets Genetics Portal analysis identified that the *MUC2* locus signal (rs779167905) allele that was associated with increased risk of chronic sputum production (allele A) was associated with higher risk of asthma and asthma-related traits in other studies [41–43] and with lower risk of gall-bladder disease (Supplementary Table 7 and 8).

#### FUT2 locus

The *FUT2* credible set included two variants that were annotated as functional using VEP. This included a stop-gain variant in *FUT2* (rs601338, linkage disequilibrium r2 0.992 with sentinel rs492602) and a nearby missense variant (rs602662 r2 0.882 with sentinel rs492602) that resulted in a Glycine to Serine amino acid change for the allele positively correlated with the chronic sputum production risk allele (Supplementary Tables 9 and 10).

Sentinel variant rs492602 at the *FUT2* locus was associated with gene expression for *FUT2, NTN5, RASIP1, SEC1P* and *MAMSTR* for which there was support for co-localisation of eQTL and GWAS signals in multiple tissues from GTEx V8 (Figure 4, Supplementary Table 11). Increased risk of chronic sputum production was consistently correlated with increased expression of *NTN5* and *MAMSTR* across a range of tissues. In contrast, the direction of the *FUT2* expression signal varied by tissue with increased risk of chronic sputum production correlated with decreased expression of *FUT2* in brain tissues and with increased expression in gastrointestinal tissue. There were no associations in lung tissue and upper airway tissues were not available.

**Figure 4.**
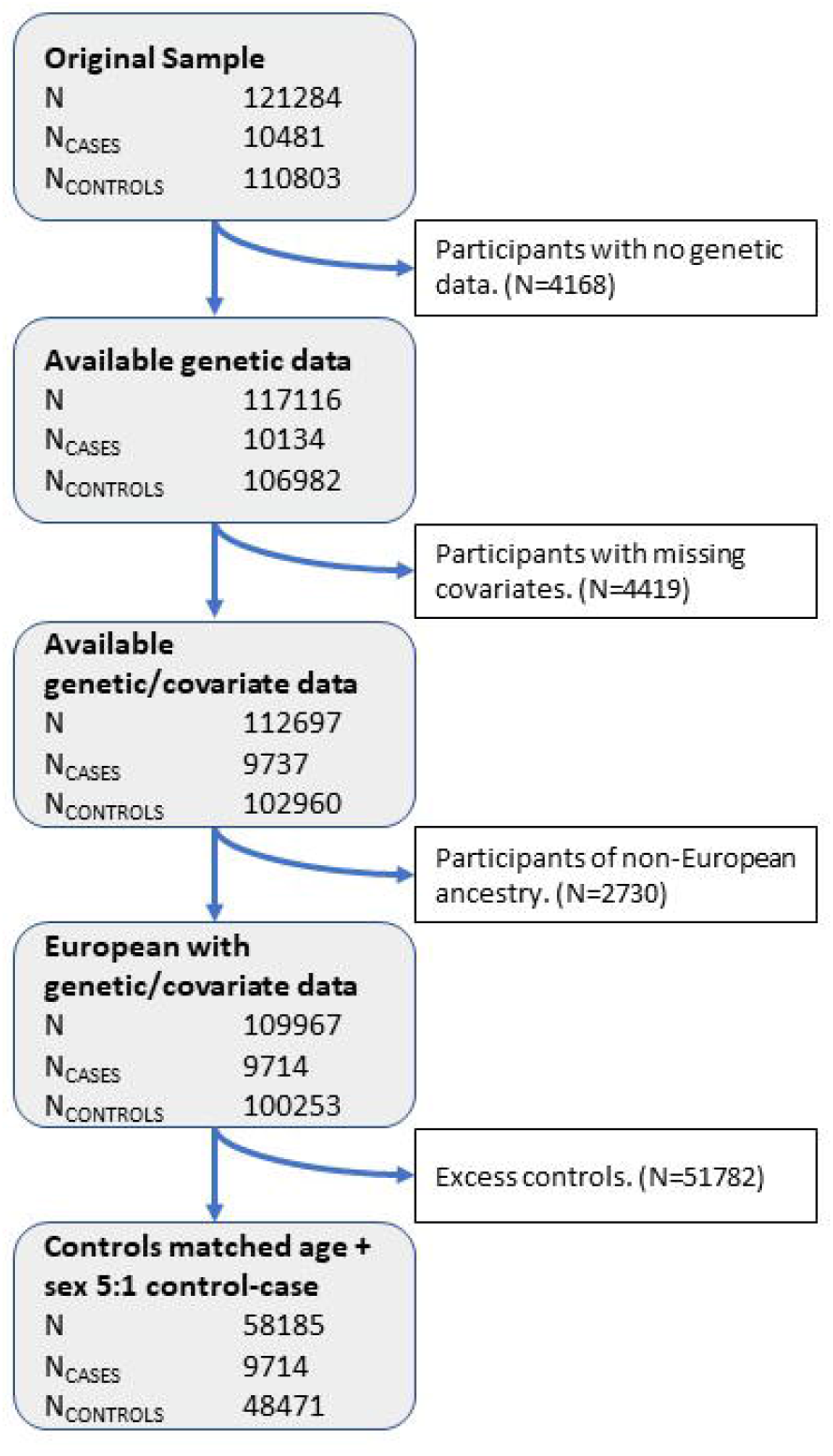
Results for eQTL colocalization for the *FUT2* locus using variant **rs492602**. The numbers within the grid are the posterior probability of colocalization (H4), with results aligned to the risk allele G for the **rs492602** variant. Missing numbers indicate no data was available for the respective gene and tissue.

The sentinel variant for the *FUT2* region signal on chromosome 19 (rs492602) was associated with lung function measures FEV_1_ /FVC and PEF (P=2.2×10^−6^ and P=1.1×10^−6^, respectively), with the chronic sputum production risk allele (G) associated with decreased lung function (Figure 3, Supplementary Table 6).

Our PheWAS and Open Targets Genetics Portal analysis for this variant identified 141 associations spanning multiple disease areas, phenotypes and biomarkers (Supplementary Tables 7 and 8). In summary, the allele associated with increased risk of chronic sputum production was associated with increased risk of gallstones [42, 44, 45], type 1 diabetes [46] and Crohn’s disease [47–50], elevated vitamin B12 [51–54] and cholesterol and fat metabolites [41, 42, 55–59], hypertension/cardiovascular disease [42, 44, 60], excess alcohol with associated sequelae [44, 61–63], increased risk of mumps and lower risk of childhood ear infections [64]. Higher risk of chronic sputum production was also associated with higher levels of gamma glutamyl transferase, total bilirubin and aspartate amino transferase, and lower levels of alanine aminotransferase and alkaline phosphatase.

#### Other novel loci

Using functional annotation of variants and eQTL analysis, no putative causal genes could be assigned to the signals in or near *OCIAD1* and *NELL1*. There was a single co-localising eQTL for *SLC25A37* in the *NKX3-1* locus with increased risk of chronic sputum production associated with a reduced expression of *SLC25A37* in brain cortex (Supplementary Table 11, Supplementary Figure 19).

## Discussion

We describe a GWAS of chronic sputum production to identify genome-wide significant signals and our novel findings implicate genes involved in mucin production and fucosylation, as well as the HLA class II histocompatibility antigen, HLA-DRB1.

The most significant signal implicated the gene *FUT2* which has been widely studied for its role in blood group antigen expression and association with gastric and respiratory infection. *FUT2* encodes fucosyltransferase 2 which mediates the transfer of fucose to the terminal galactose on glycan chains of cell surface glycoproteins and glycolipids. FUT2 creates a soluble precursor oligosaccharide FuC-alpha ((1,2)Galbeta-) called the H antigen which is an essential substrate for the final step in the soluble ABO blood group antigen synthesis pathway. The *FUT2* locus allele associated with increased risk of chronic sputum production in this study is correlated with a nonsense allele that leads to inactivated FUT2, which results in a non-secretory phenotype of ABO(H) blood group antigens [65] for homozygous carriers. This nonsense allele (rs601338 allele A) has frequencies of 25-50% in South Asian, European and African populations but is rare (<1%) in East Asian populations [66]. Candidate gene studies of this locus have identified that non-secretors (at increased risk of chronic sputum production according to our study) have a lower risk of H. Pylori infection [67], rotavirus A infection [68, 69], norovirus infection [70–72], infant (12-24 months) respiratory illness [73], asthma exacerbations [74], otitis media [75], exacerbation in non-cystic fibrosis bronchiectasis and *Pseudomonas aeruginosa* airway infection in the same group [76], some evidence of slower HIV progression [72] and a higher risk of pneumococcal and meningococcal infection [77]. The T allele of another variant in high linkage disequilibrium at this locus (rs681343, r^2^=0.996 with rs492602), associated with increased risk of chronic sputum production in our study, was recently reported to be associated with increased risk of human polyomavirus 1 (BKV) virus infection, as measured by antibody response [78]. A recent GWAS of critically ill cases of COVID-19 (cases N=7491), showed that the risk allele for chronic mucus production (G) of rs492602 was protective against life threating COVID-19 (P=4.55×10^−9^, OR 0.88, CI 0.87-0.90) [79]. However, this finding was not replicated in the latest COVID-19 Host Genetics Initiative results for a similar phenotype [80]. The differing directions of effect of this signal on different phenotypes may be explained by the SNP effects on FUT2 expression which differ across cell and tissue types. Further targeted experiments in relevant cell and tissue types would be needed to elucidate this and define the likely effects of targeting FUT2 directly or indirectly.

Epitopes that are fucosylated by FUT2 play a role in cell-cell interaction including host-microbe interaction [81, 82] and mediate interaction with intestinal microbiota, thereby influencing its composition [83–86]. Whilst there has been no direct evidence of host-pathogen binding on the FUT2 generated epitopes for non-gastrointestinal infection there is evidence that FUT2 can influence non-binding ligands such as sialic acid [87]. Sialic acid binding has been shown to be important for adenovirus binding in cell models [88] and modulating this binding has been implicated as a possible mechanism for increasing risk of mumps infection [64].

FUT2 may also be key to the function of mucins, including those encoded by genes at our other significant locus (i.e. *MUC2, MUC5AC, MUC5B*). Analysis of oligosaccharides released from insoluble colonic mucins, largely Muc2, by mass spectrometry shows complete lack of terminal fucosylation of *O-*linked oligosaccharides in Fut2-LacZ-null mice [89]. FUT2 has also been shown to determine the *O-* glycosylation pattern of Muc5ac in mice [90]. The significant signal at *MUC2* in our analysis was not independent of the previously reported moderate-to-severe asthma signal [20] for which *MUC5AC* was implicated as the most likely causal gene using gene expression data from bronchial epithelial cells. Although our analysis did not identify an association at the *MUC2* locus with COPD-related traits (FEV_1_ and FEV_1_/FVC), a recent study has also highlighted MUC5AC as a potential biomarker for COPD prognosis [91].

The particular allele that was found to explain the association signal in the HLA region (HLA-DRB1*03:147 [92], has only recently been reported and so there is limited information about functionality. Associations of this allele with other GWAS loci should be interpreted with caution given the high LD across the region.

Through identification of genetic association signals that are independent of smoking and history of chronic respiratory disease, our study demonstrates the value in studying a disease-relevant phenotype in a very large population that is agnostic to respiratory disease or smoking status. We only report overlap of chronic sputum production association signals with association signals for gene expression regulation where there is statistical support that these signals share a causal variant. In addition to a comprehensive PheWAS, we provide a deeper assessment of associations with relevant respiratory phenotypes that highlights previously unreported associations with lung function for the *HLA-DRB1* and *FUT2* signals.

As only a subset of UK Biobank participants provided answers to the sputum production question, we expected that we might be able to define a replication case control dataset from the remaining >300,000 participants using primary care data. However, evaluation of the positive predictive value of primary care codes for sputum production, when compared to the questionnaire data, was very low (see Supplementary Text). This could reflect a low utilisation of sputum codes in primary care or that participants have not reported this symptom to their General Practitioner (GP). We obtained supportive evidence for four of the signals utilising data from five general population cohorts. The limited sample size (the case sample size for replication was 23% of the size available for discovery) impacted our ability to show statistically significant replication. Furthermore, we note that, for three of the replication cohorts (LIfeLines 1 and 2 and Vlagtwedde-Vlaardingen), the sputum production question asked specifically about winter symptoms whilst the UK Biobank survey did not restrict to any specific season. However, given the strong evidence summarised above for the involvement of the probable causal genes in control of pathways relevant to mucus production, we believe the associations identified are highly likely to be real. Due to very low numbers, we were unable to evaluate the effects of these signals in individuals of non-European ancestry thereby limiting the generalisability of our findings to non-European ancestry groups. Efforts are urgently needed to improve diversity in genomics research [93] such as the planned Our Future Health initiative in the UK.

In summary, the HLA, *MUC2* and *FUT2* loci show strong candidacy for a role in sputum production, with overlap with infection and related phenotypes and known mechanistic interactions between the genes at the *FUT2* and *MUC2* loci, suggesting that these signals are likely to be robust. The large number of associations of the *FUT2* locus with a broad array of phenotypes, tissue-dependent expression of *FUT2*, and association with expression of other genes in the region, may have implications for drug targeting guided by this locus. Experimental studies to characterise the specific interplay between FUT2 activity and mucin genes expressed in the airways are warranted.

## Conclusion

Chronic sputum production is a phenotype characteristic of several respiratory diseases, as well as being common cause for referrals in the absence of overt disease, and is of interest for pharmaceutical intervention. We report novel genetic factors which influence chronic sputum production and these signals highlight fucosylation of mucin as a driving factor of chronic sputum production. These signals could provide insight into the molecular pathways of sputum production and represent potential future targets for drug development [94].

## Supporting information

STEGA Strobe checklist

## Data Availability

Genome-wide association statistics from the case-control analysis of chronic sputum production will be made available via GWAS Catalog [to be submitted following peer-review].

## Funding and Acknowledgements

L.V.W. holds a GSK / Asthma + Lung UK Chair in Respiratory Research (C17-1). M.D.T. is supported by a Wellcome Trust Investigator Award (WT202849/Z/16/Z). M.D.T. and I.P.H. hold NIHR Senior Investigator Awards. C.J. held a Medical Research Council Clinical Research Training Fellowship (MR/P00167X/1). LVW, MDT, IS and IPH report collaborative research funding from GSK to undertake the submitted work.

The research was partially supported by the NIHR Leicester Biomedical Research Centre and the NIHR Nottingham Biomedical Research Centre; the views expressed are those of the author(s) and not necessarily those of the NHS, the NIHR or the Department of Health. This research was funded in part by the Wellcome Trust. We acknowledge the support of the Health Data Research UK BREATHE Digital Innovation Hub (UKRI Award MC_PC_19004). This research was conducted under UK Biobank application 45243. This research used the SPECTRE and ALICE High Performance Computing Facility at the University of Leicester.

Generation Scotland received core support from the Chief Scientist Office of the Scottish Government Health Directorates [CZD/16/6] and the Scottish Funding Council [HR03006] and is currently supported by the Wellcome Trust [216767/Z/19/Z]. Genotyping of the GS:SFHS samples was carried out by the Genetics Core Laboratory at the Edinburgh Clinical Research Facility, University of Edinburgh, Scotland and was funded by the Medical Research Council UK and the Wellcome Trust (Wellcome Trust Strategic Award “STratifying Resilience and Depression Longitudinally” (STRADL) Reference 104036/Z/14/Z). CH is supported by a Medical Research Council University Unit Programme grant MC_UU_00007/10 (QTL in Health and Disease).

Recruitment to the Generation Scotland CovidLife study was facilitated by SHARE - the Scottish Health Research Register and Biobank.

SHARE is supported by NHS Research Scotland, the Universities of Scotland and the Chief Scientist Office of the Scottish Government.

For the purpose of open access, the author has applied a CC BY public copyright licence to any Author Accepted Manuscript version arising from this submission.

