## Supplementary Methods and Figures for "Genome-wide association study of chronic sputum production implicates loci involved in mucus production and infection"

Supplementary Materials

### Supplementary Text

#### Replication: Description of independent replication cohorts (Generation Scotland, LifeLines 1, LifeLines 2, Vlagtwedde—Vlaardingen and EXCEED Study)

##### The Generation Scotland study

The Generation Scotland study (GS) is a population- and family-based cohort with broad consent for genetic, health, well-being and lifestyle studies. (Smith *et al.*, 2013) The main recruitment (24,096 individuals in 5501 family groups) took place during 2006–11.

In 2020, a series of CovidLife surveys were conducted during the COVID-19 pandemic. Survey invitations were sent to 22,796 members of GS who provided an e-mail address for recontact, as well as to other adults in the UK through collaborators and social media channels (Fawns-Ritchie *et al.*, 2021).

##### LifeLines 1 and 2 and Vlagtwedde-Vlaardingen

Genotyped individuals from the first (n = 7,976) and second (n = 5,260) data release of the LifeLines cohort study (2006–2011) (Scholtens *et al.*, 2015) and 1,529 subjects from the last survey (1989/1990) from the Vlagtwedde-Vlaardingen cohort (de Jong *et al.*, 2014; van Diemen *et al.*, 2005), a prospective general population based cohort including Caucasians of Dutch descent were used as replication cohorts (Zeng *et al.*, 2017).

In these cohorts, genotyping was performed using IlluminaCytoSNP-12 arrays. The applied genotyping quality control criteria have been described before (de Jong *et al.*, 2015; Scholtens *et al.*, 2015): Samples with call-rates of less than 95% were excluded as were samples of non-Caucasians and first degree relatives. SNPs were excluded if they had a genotype call-rate < 95%, minor allele frequency (MAF) < 1%, or a Hardy-Weinberg equilibrium (HWE) p-value < 10−4.

Phlegm was measured by standardized questionnaires from the European Community Respiratory Health Survey (ECRHS) (Burney *et al.*, 1994). Phlegm was defined as at least one positive answer to the questions: “do you usually bring up any phlegm from your chest first thing in the morning in winter?” or “do you usually bring up any phlegm from your chest during the day, or at night, in winter?”.

The analysis on the presence of phlegm was performed using PLINK version 1.07 (Purcell *et al.*, 2007). We used an additive genetic model adjusted the logistic regression analysis for age, sex, and current smoking.

##### EXCEED

EXCEED is a longitudinal population-based cohort which facilitates investigation of genetic, environmental and lifestyle-related determinants of a broad range of diseases and of multiple morbidity through data collected at baseline and via electronic healthcare record linkage. Recruitment has taken place in Leicester, Leicestershire and Rutland since 2013 and is ongoing, with 11,000 participants. Recruitment was widened to anyone over 18 years of age living in the Midlands in 2020. Participants provided a DNA sample, have consented to follow-up for up to 25 years through electronic health records and additional bespoke data collection is planned. Data available includes baseline demographics, anthropometry, spirometry, lifestyle factors (smoking and alcohol use) and longitudinal health information from primary care records, with additional linkage to other EHR datasets planned. Patients have consented to be contacted for recall-by-genotype and recall-by-phenotype sub-studies. Further details about the study can be accessed in the Cohort Profile Paper(John *et al.*, 2019), with additional information about our COVID-19 Focus available as a Data Note Paper (Lee *et al.*, 2021).

#### Replication: Defining sputum phenotype using primary care data in UK Biobank

We evaluated whether primary care codes for sputum production could be used to define an independent case-control dataset within UK Biobank for replication of our discovery GWAS. To do this, we first evaluated the overlap of Read v3 codes for sputum (Supplementary Table 12) in the available V1.0 September 2019 primary care data for the 121,283 participants who had answered “yes” or “no” for UK Biobank field 22504 (“do you bring up phlegm/sputum/mucus daily?”). To be deemed useful to identify new cases from the remaining participants with primary care data, we required a positive predictive value to predict a case based on primary care records to be above 80%.

Presence of one or more sputum codes had a Positive Predictive Value (PPV) of 29% (7.9% of cases and 1.9% of controls had one or more Read codes indicative of sputum production). This is unlikely to change with the release of additional UK Biobank primary care data and this we concluded that we were unable to use primary care data in UK Biobank to define an independent replication dataset.

#### Associations with other phenotypes: chronic cough and chronic bronchitis

We defined four phenotypes using UK Biobank data for association analysis with the sentinel variants. We defined cough and chronic bronchitis phenotypes selecting cases as those answering yes and controls answering no to the following UK Biobank data-fields, 22502 (cough) 22124 (chronic bronchitis). All phenotypes were limited to European ancestry using the same methods as the primary analysis, association with cough and chronic bronchitis used the same covariates as the primary analysis, all association analyses were run in PLINK 2.0 (Chang et al., 2015).

#### Associations with other phenotypes: PheWAS analysis

Traits included UK Biobank baseline measures (from questionnaires and physical measures), self-reported medication usage, and operative procedures, as well as those captured in Office of Population Censuses and Surveys codes from the electronic health record. We also included self-reported disease variables and those from hospital episode statistics (ICD-10 codes truncated to three-character codes and combined in block and chapter groups), combining these where possible to maximize power. A total of 2,150 traits were defined with >200 cases and were included for analysis. Analyses were conducted in unrelated European-ancestry individuals (KING kinship coefficient < 0.0442), and were adjusted for age, sex, genotyping array and first ten principal components. Logistic and linear models were fitted for binary and quantitative outcomes, respectively. Biomarker measurements were adjusted for statins according to the ‘Statin identification and LDL adjustment’ methods described by Sinnott-Armstrong 2019 (Sinnott-Armstrong *et al.*, 2021).  Statin-adjusted phenotype values were further adjusted (in residualization) for age at assessment center visit, genetic sex, genotyping array, fasting time, sample dilution factor, socio-economic status indicator, blood sample hour, and urine sample hour with assessment center as a random effect.  We then conducted rank-based inverse normal transformation of these residuals.  The rank-based inverse-normal transformed residuals were used as inputs for our GWAS linear regression in Hail. False discovery rate (FDR) was calculated using the Benjamin H method (Benjamini and Hochberg, 1995) adjusting for the 2,172 traits tested. Associations with a FDR <0.05 are reported.

To test association with the HLA allele we used, DeepPheWAS (Packer *et al.*, 2022). The platform includes 2,504 phenotypes in UK Biobank, a subset of 2,246 are recommended for association testing. Deep PheWAS then filters these requiring a minimum case number, we chose to keep the default settings of 50 case minimum for binary phenotypes and a 100-case minimum for quantitative phenotypes. After limiting to European ancestry, filtering for case numbers and removing related pairs (KING kinship coefficient >=0.0884) this left 1,907 phenotypes for association analysis.

### Supplementary Figures

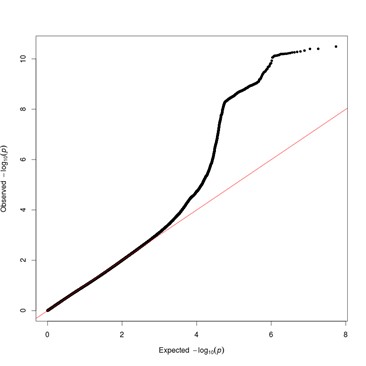

**Supplementary Figure** 1: Quantile-quantile (QQ) plot for results of genome-wide association study of chronic sputum production in UK Biobank. Variants with imputation quality INFO <0.5 and minor allele count (MAC) <20 were excluded. The genomic control inflation factor, lambda, was 1.026.

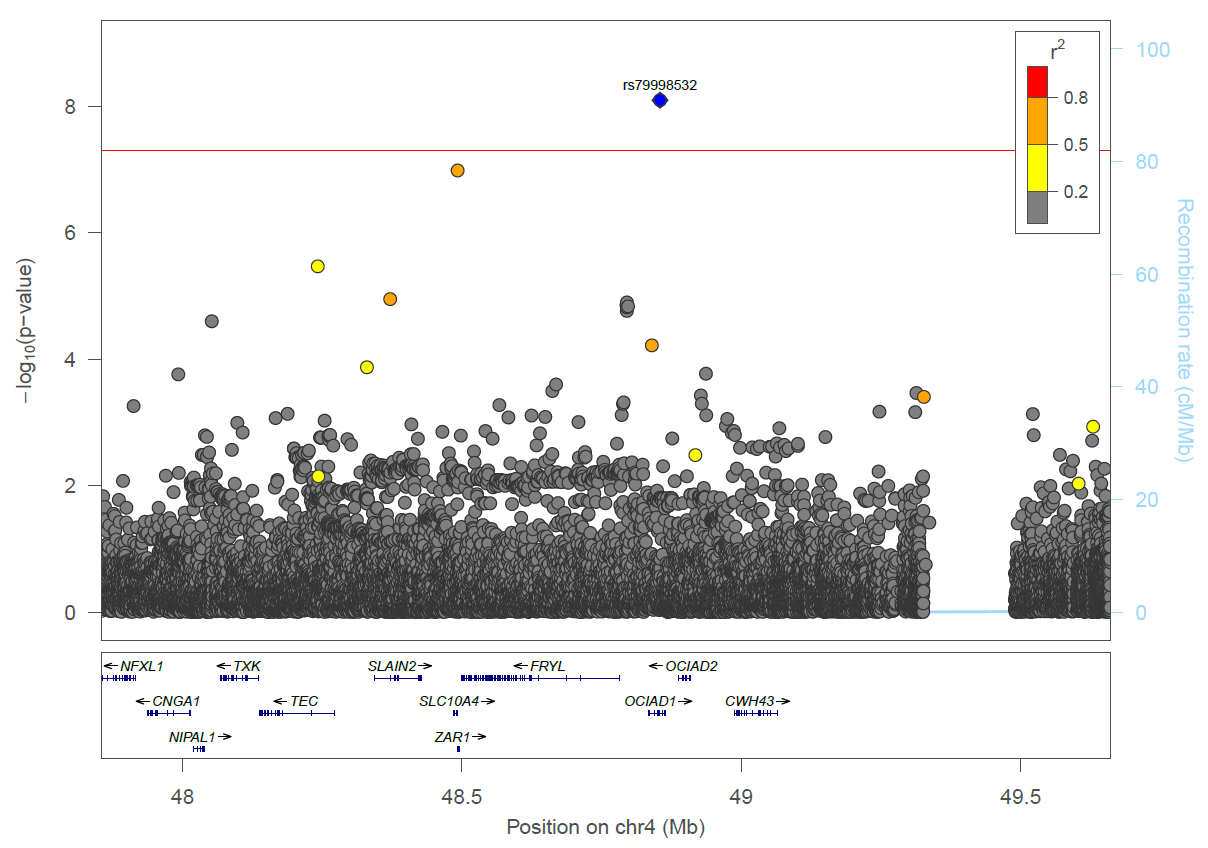

**Supplementary Figure 2:** *OCIAD1* signal (rs79998532) regional association plot.

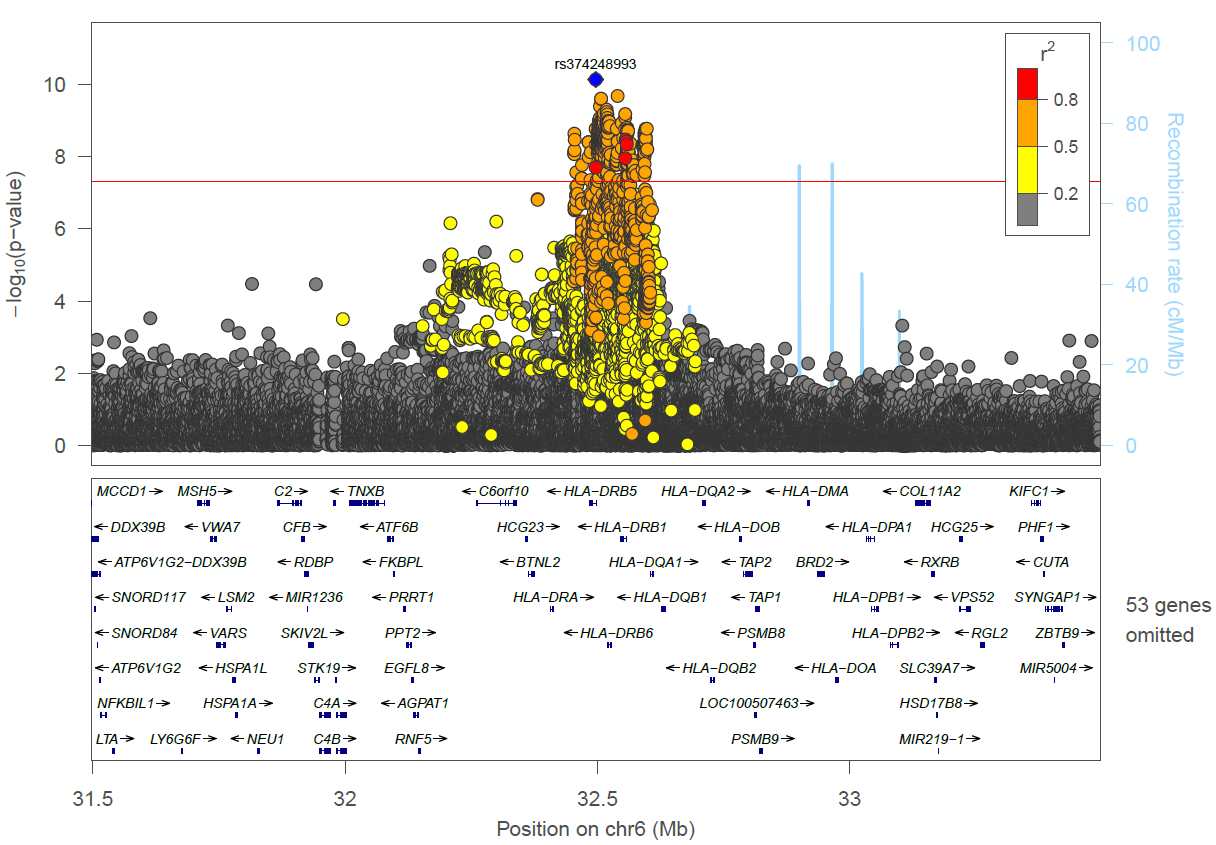

**Supplementary Figure 3:** *HLA-DRB5* signal (rs374248993) regional association plot.

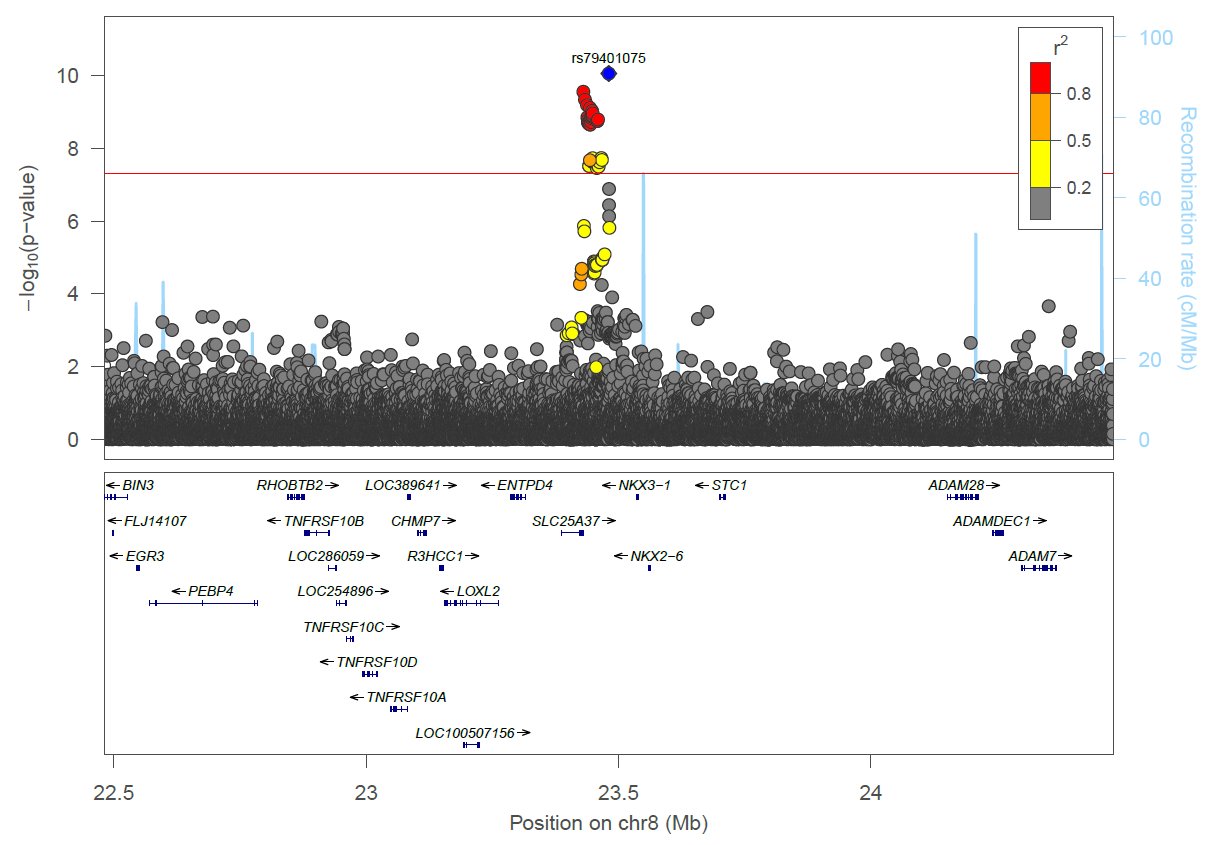

**Supplementary Figure 4:** *NKX3-1* signal (rs79401075) LocusZoom plot.

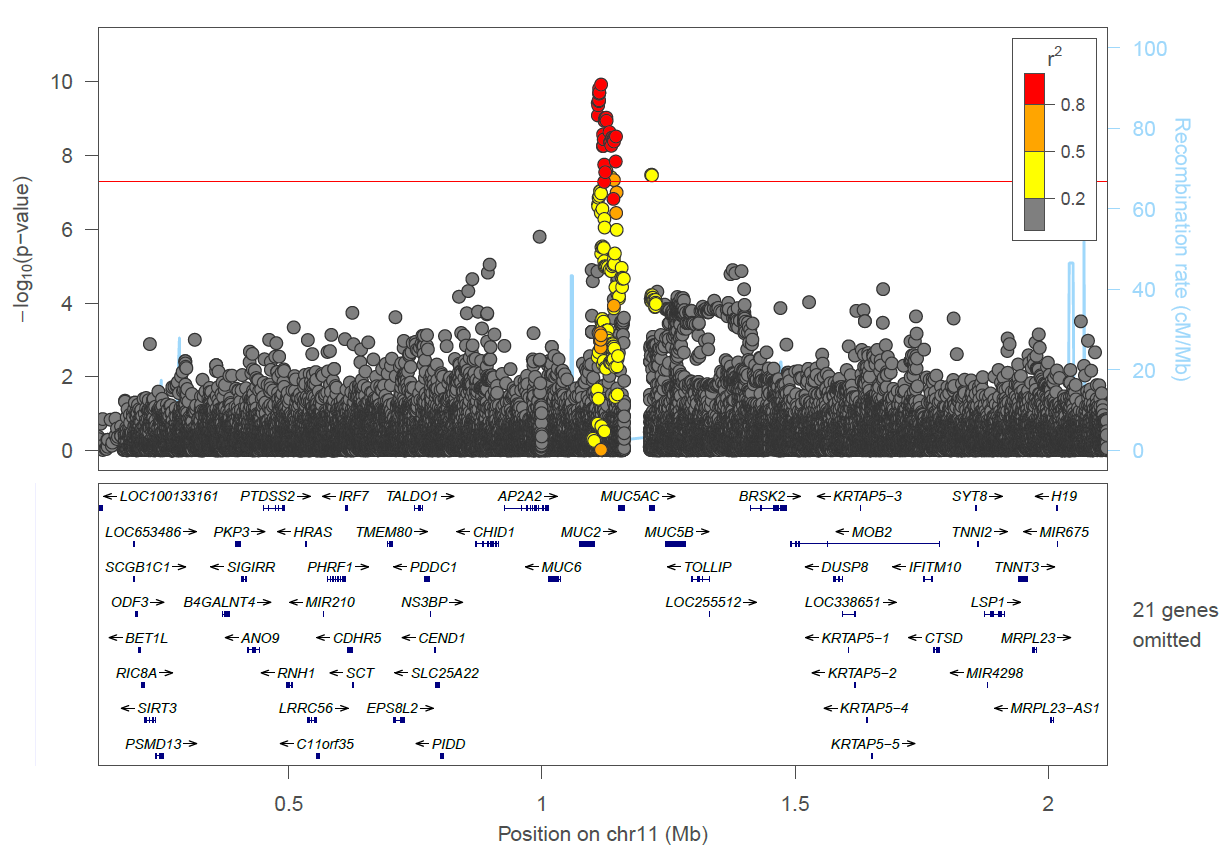

**Supplementary Figure** **5:** *MUC2* signal (rs779167905) regional association plot.

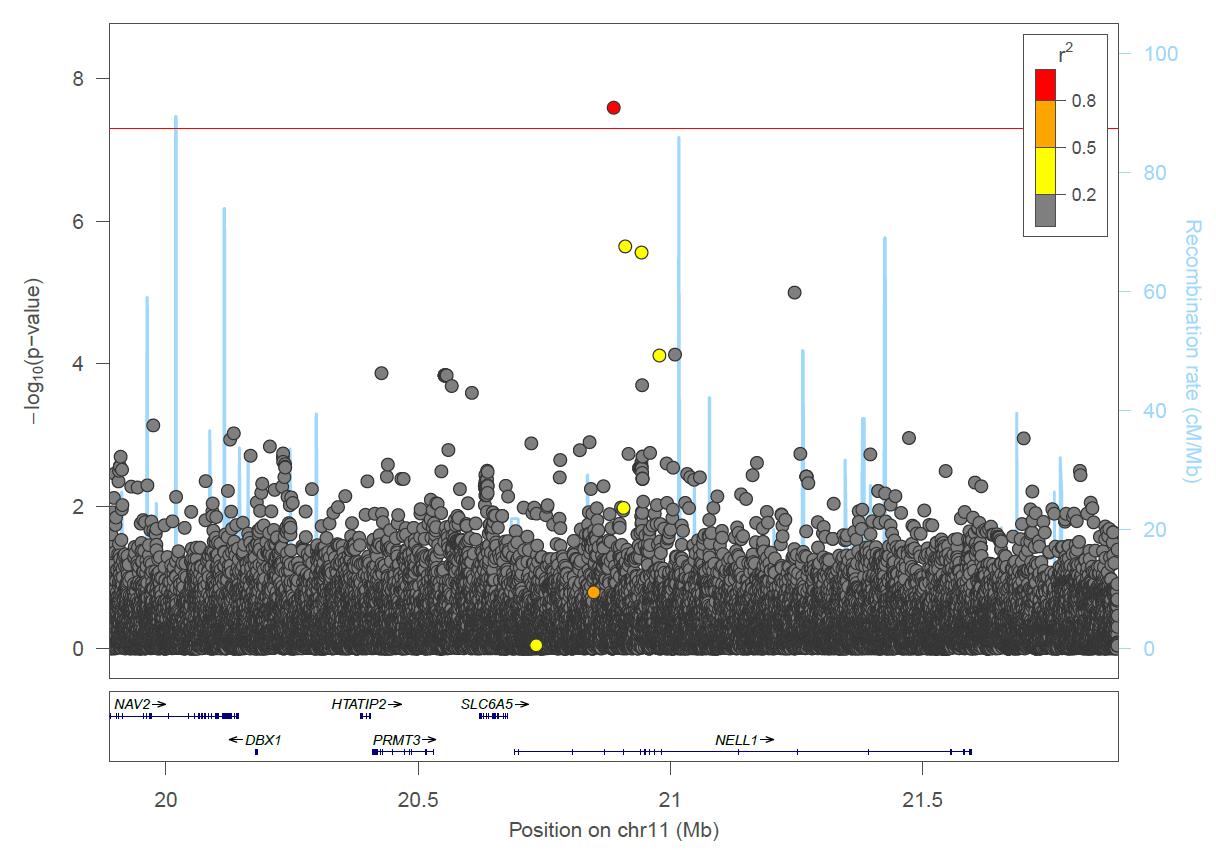

**Supplementary Figure 6:** *NELL1* signal (rs529240826) regional association plot.

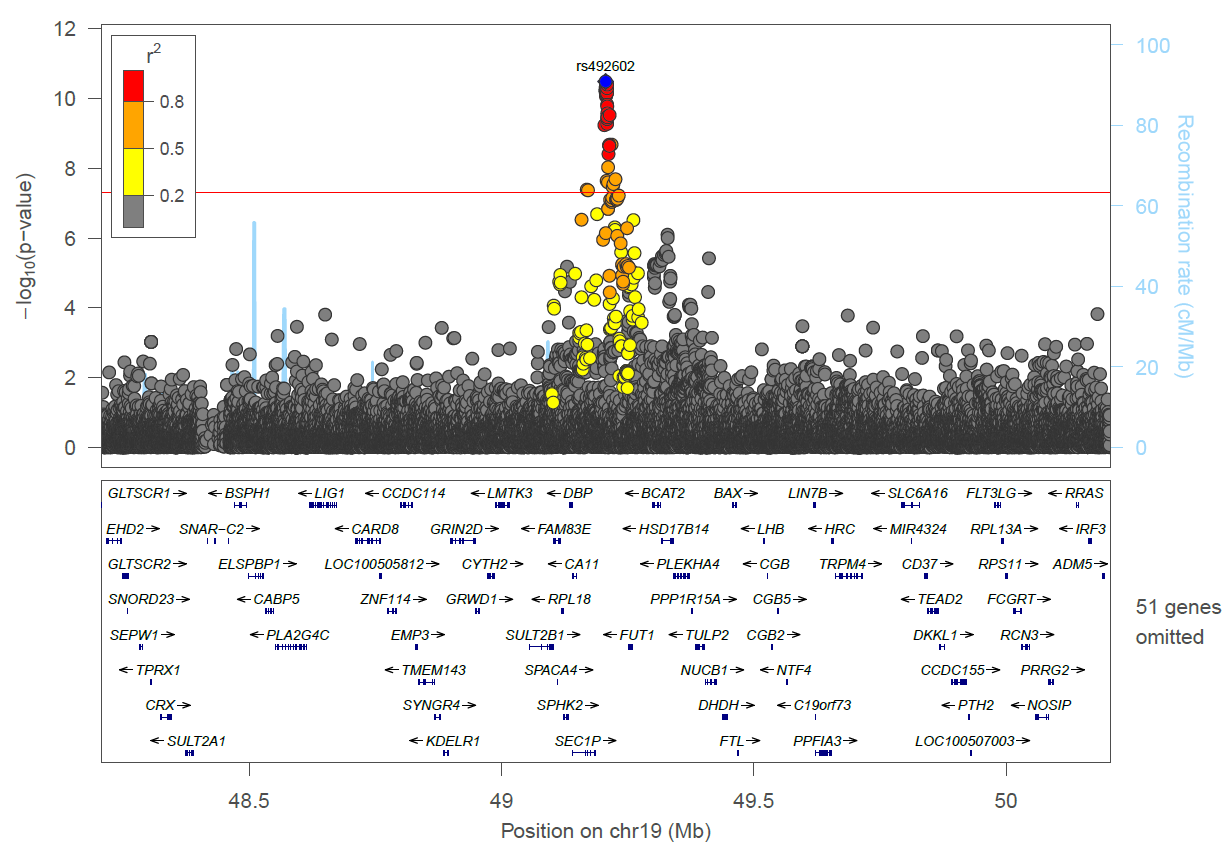

**Supplementary Figure 7**: *FUT2* signal (rs492602) regional association plot.

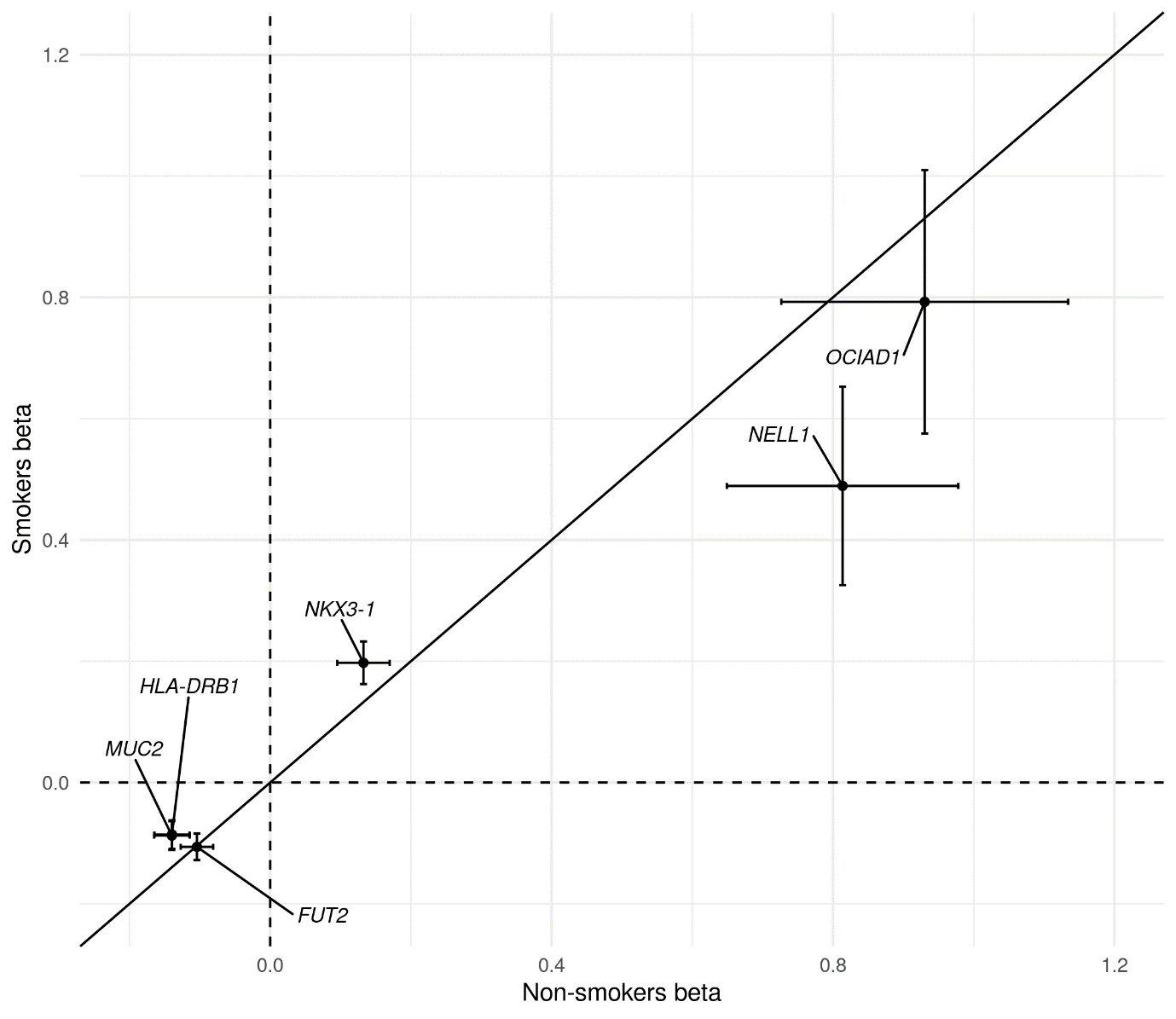

**Supplementary Figure 8:** Plot of beta values for association for sentinel variants in smokers (N cases = 5,306, N controls = 20,913) against beta values for sentinel variants in non-smokers (N cases = 4,408, N controls = 27,559), sentinel variants labelled with corresponding loci.

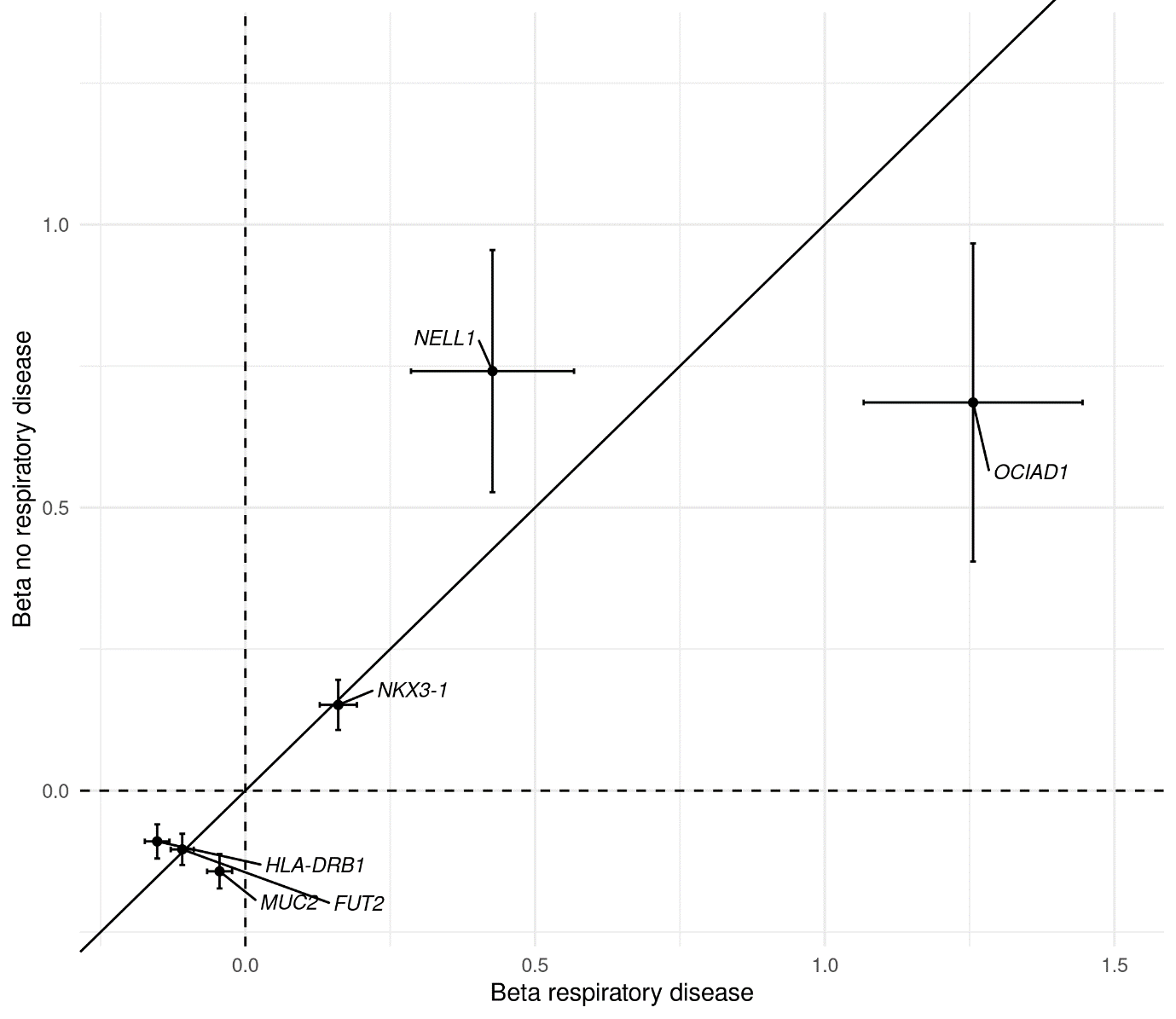

**Supplementary Figure 9:** Plot of beta values for association for sentinel variants in those with no history of chronic respiratory disease (N cases = 39,967, N controls = 6,076), against beta values for sentinel variants in those with a history of chronic respiratory disease (N cases = 3,741 , N controls = 9,118), sentinel variants labelled with corresponding loci. History of respiratory disease defined as one or more of, spirometry defined COPD GOLD1+, doctor diagnosed asthma or doctor diagnosed chronic bronchitis.

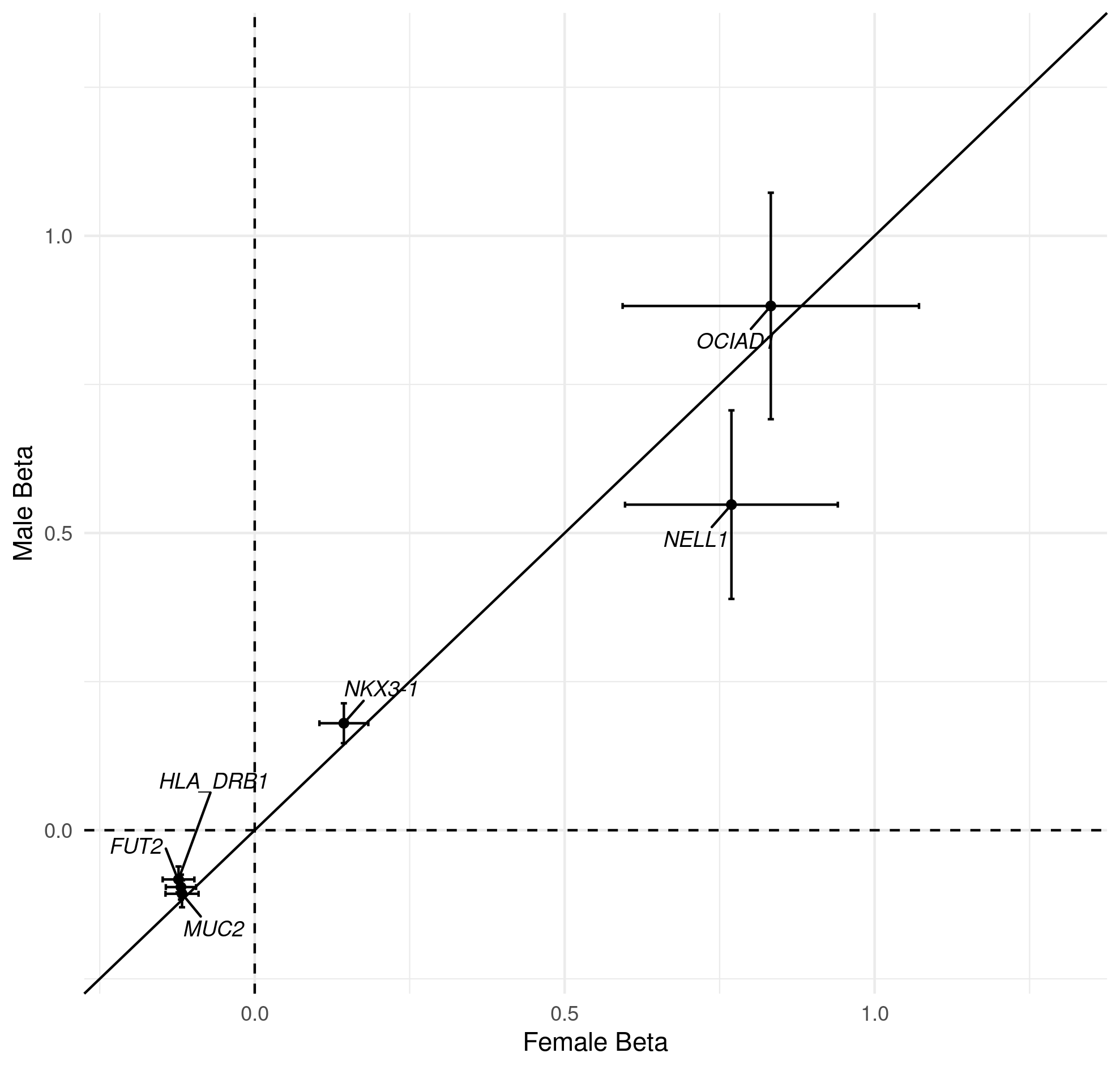

**Supplementary Figure 10:** Plot of beta values for association for sentinel variants in males (N cases = 5,589, N controls = 27,880), against beta values for sentinel variants in females (N cases = 4,124, N controls = 20,579), sentinel variants labelled with corresponding loci.

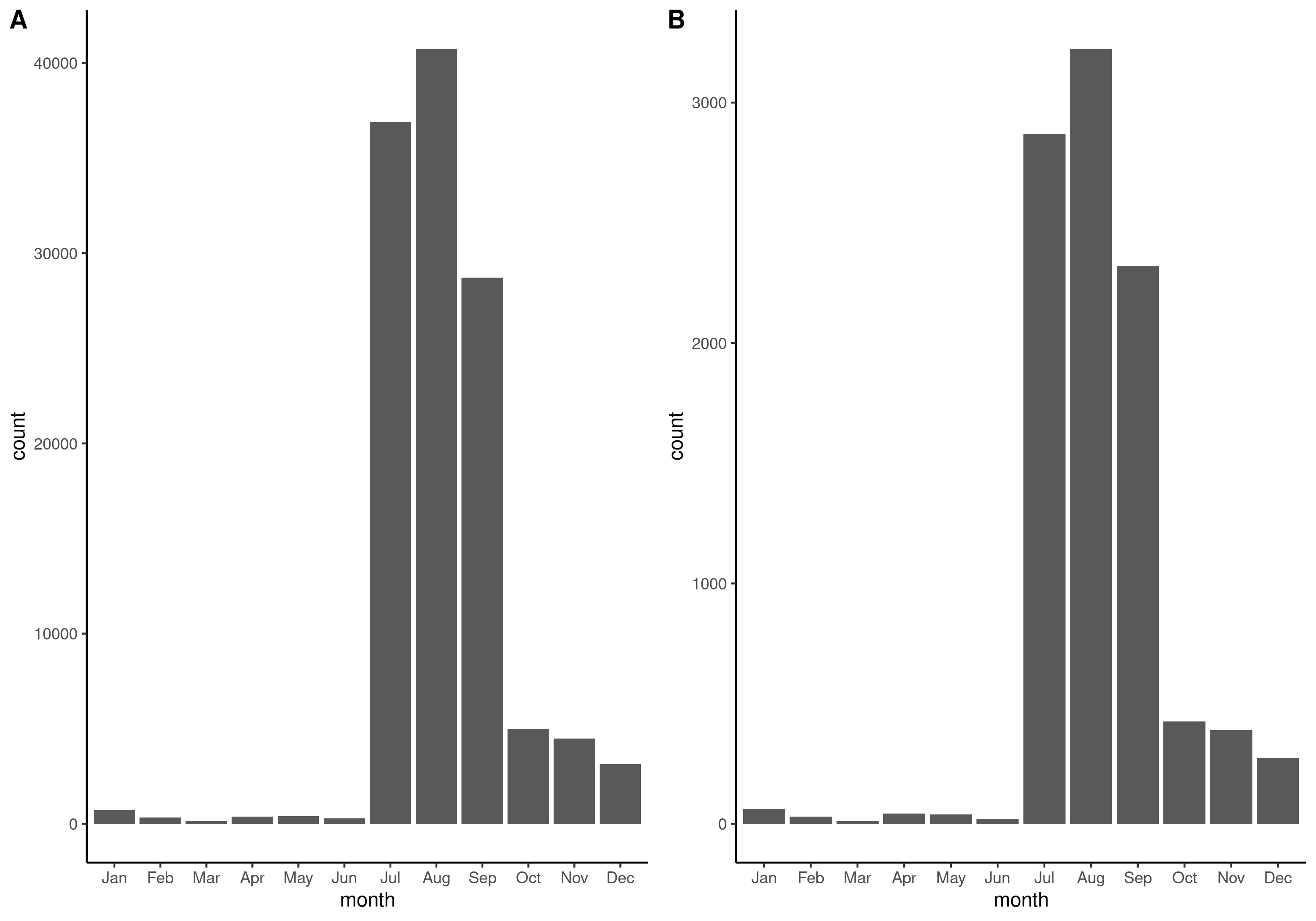

**Supplementary Figure 11:** Histogram of month that online questionnaire was completed (field-ID 22500) for A) all cases and controls and B) cases only in UK Biobank.

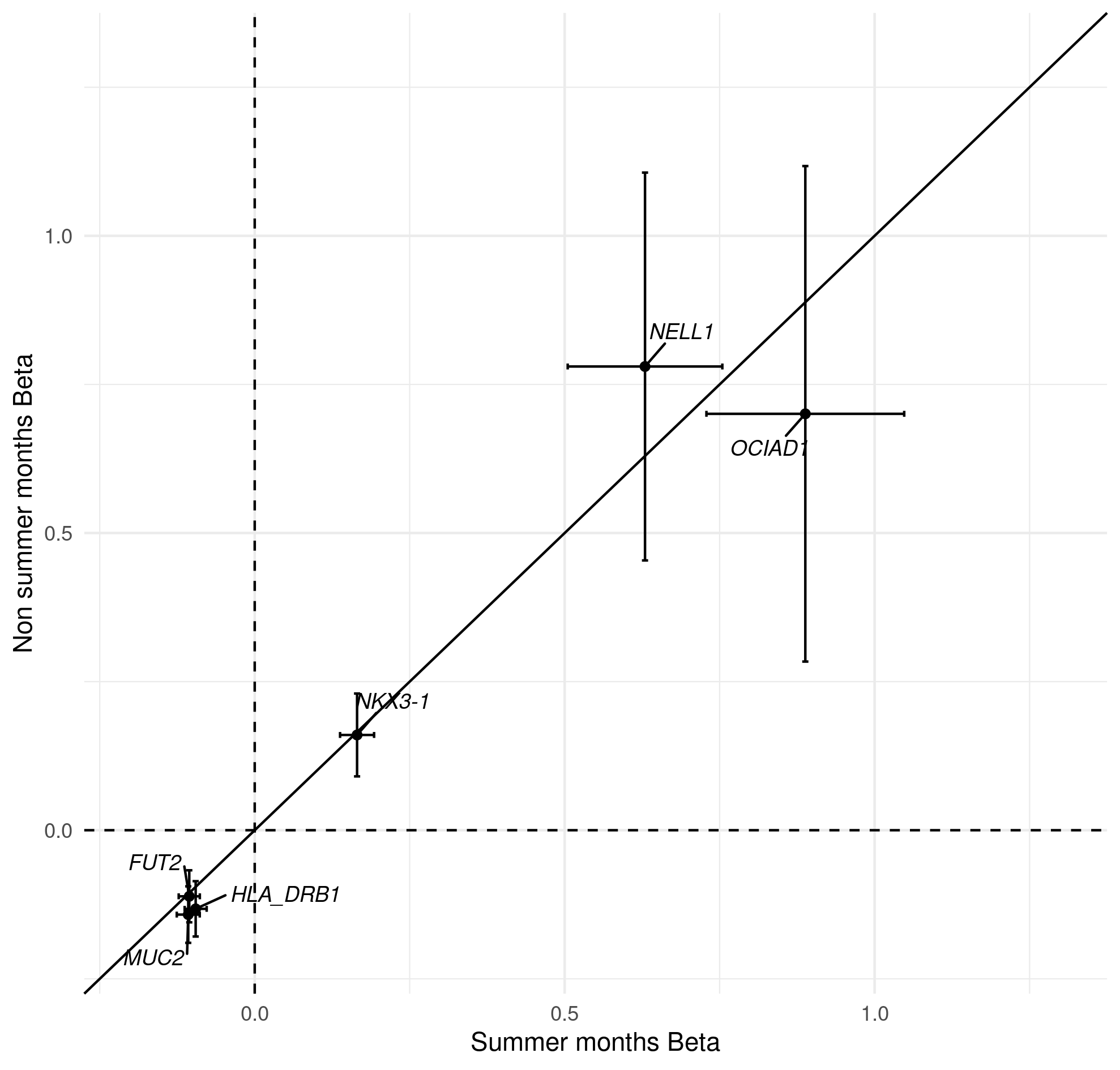

**Supplementary Figure** **12:** Plot of beta values for association for sentinel variants in those who completed the online questionnaire in July, August, and September (N cases = 8,416, N controls = 42,627), against beta values for sentinel variants in those who completed the online questionnaire in the other months (N cases = 1,297, N controls = 5,832) sentinel variants labelled with corresponding loci. July, August, and September selected as these contained the most correspondents in months with higher allergen exposure, see Supplementary Figure 11.

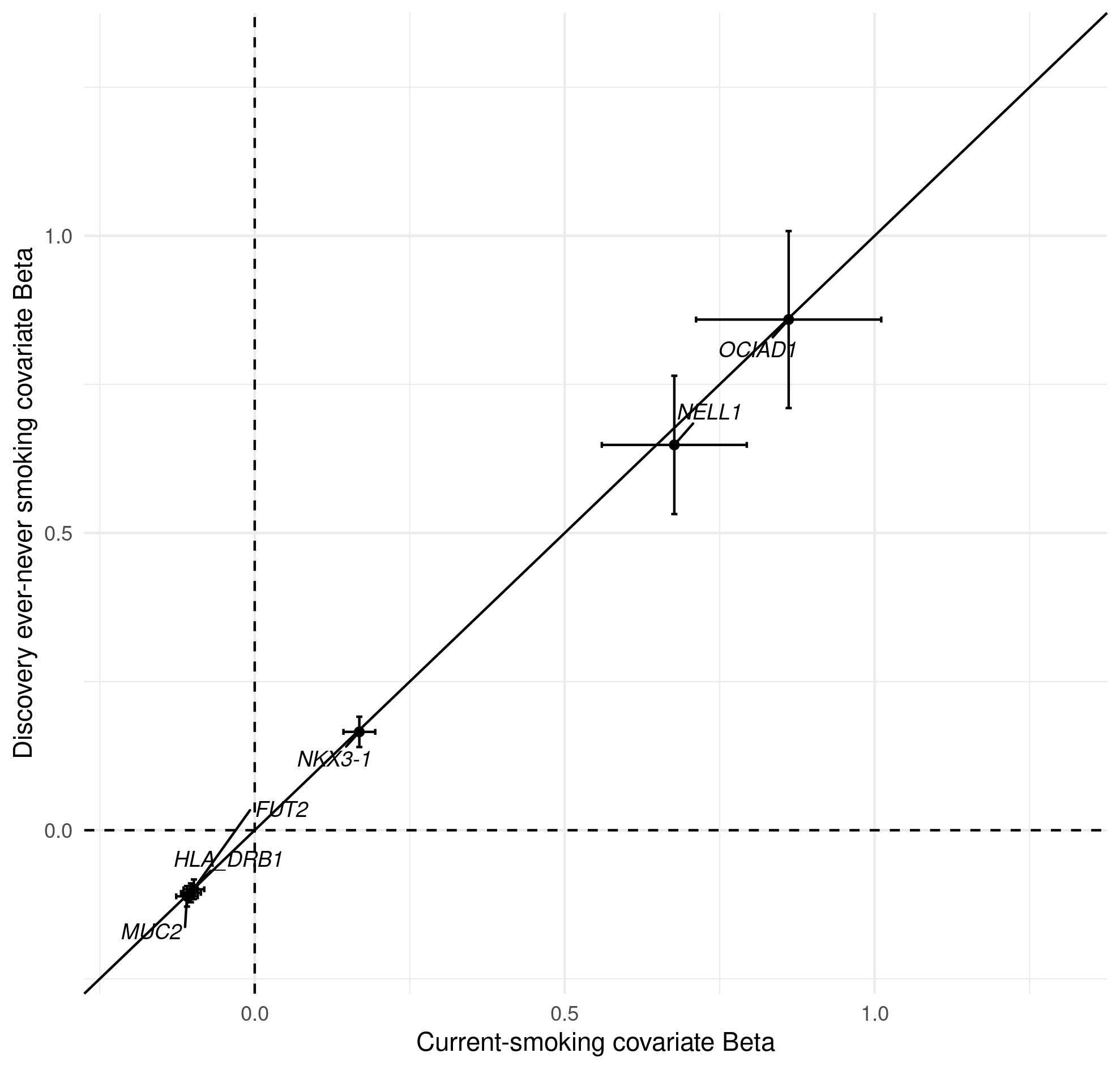

**Supplementary Figure** **13:** Plot of beta values for association for sentinel variants with covariate ‘current smoker’ replacing ‘ever smoker’ in discovery sample (N cases = 9,714, N controls = 48,471), against beta values for sentinel variants in the discovery with original covariates (N cases = 9,714, N controls = 48,471), sentinel variants labelled with corresponding loci.

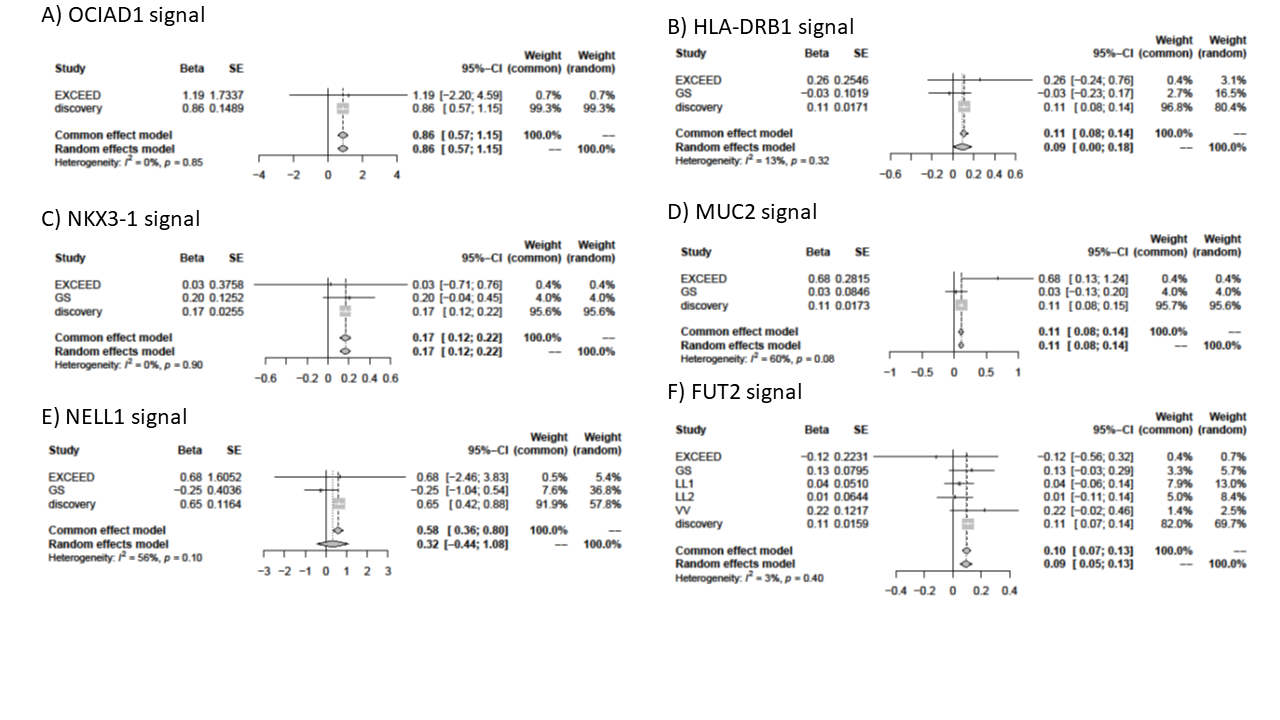

**Supplementary Figure 14** Forest plots for results from discovery and replication studies. GS: Generation Scotland, LL1: LifeLInes 1, LL2: LifeLines 2, VV: Vlagtwedde-Vlaardingen.

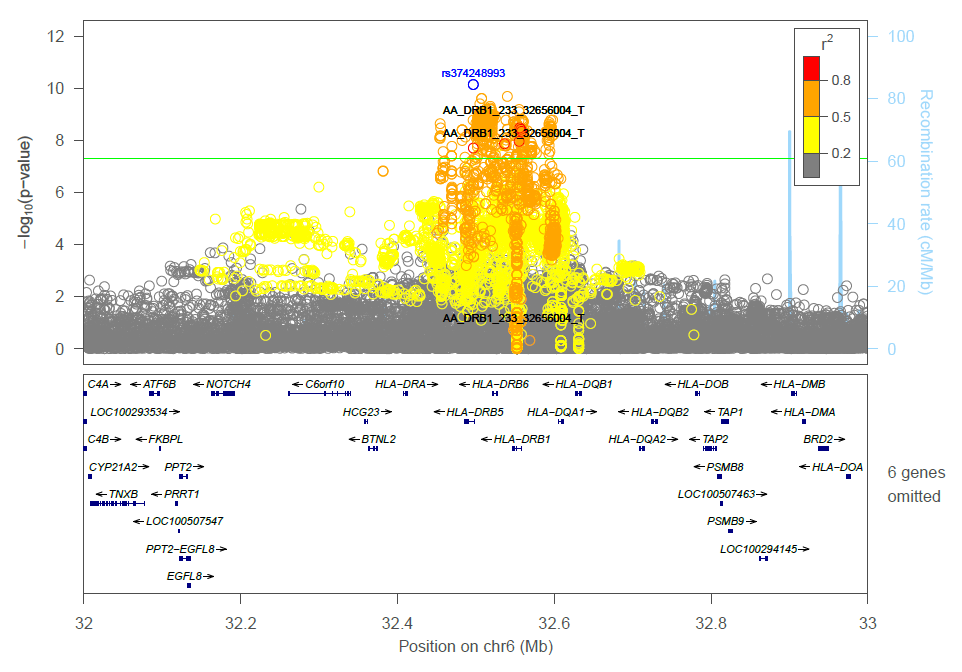

**Supplementary Figure 15**: Locuszoom plot for combined results SNPs, HLA allele and amino acid changes for *HLA-DRB1* locus, HLA alleles have been labelled in black, sentinel variant is labelled and colored blue.

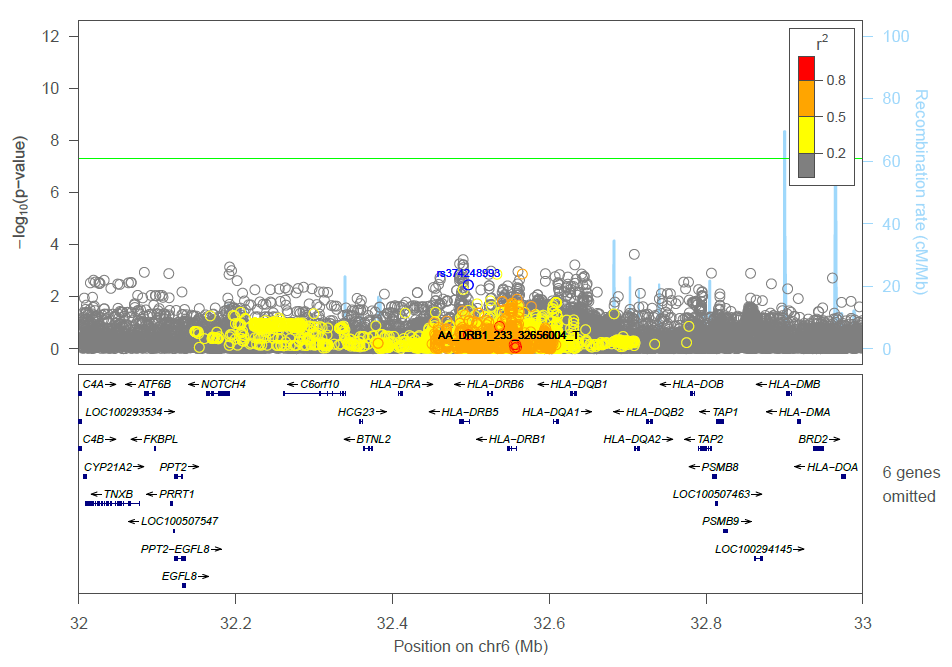

**Supplementary Figure 16:** Locuszoom plot for combined results SNPs, HLA allele and amino acid changes for *HLA-DRB1* locus conditioned on HLA allele AA_DRB1_233_32656004_T (*DRB1**03:147), HLA alleles have been labelled in black, sentinel variant is labelled and colored blue

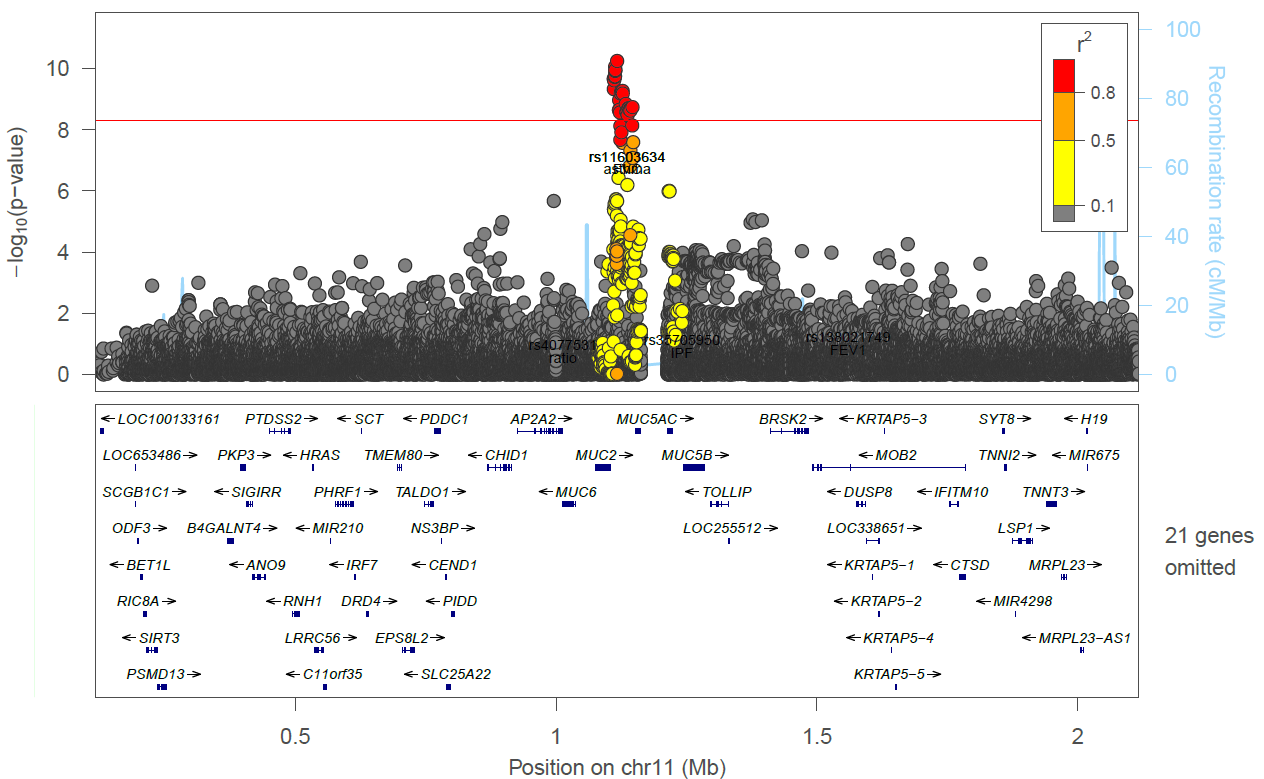

**Supplementary Figure 17:** Region plot of **rs779167905 signal** conditioned on IPF signal **rs7934606** (r^2^ = 0.005 with chronic sputum sentinel rs779167905, after conditioning chronic sputum P = 5.67×10^-11^)

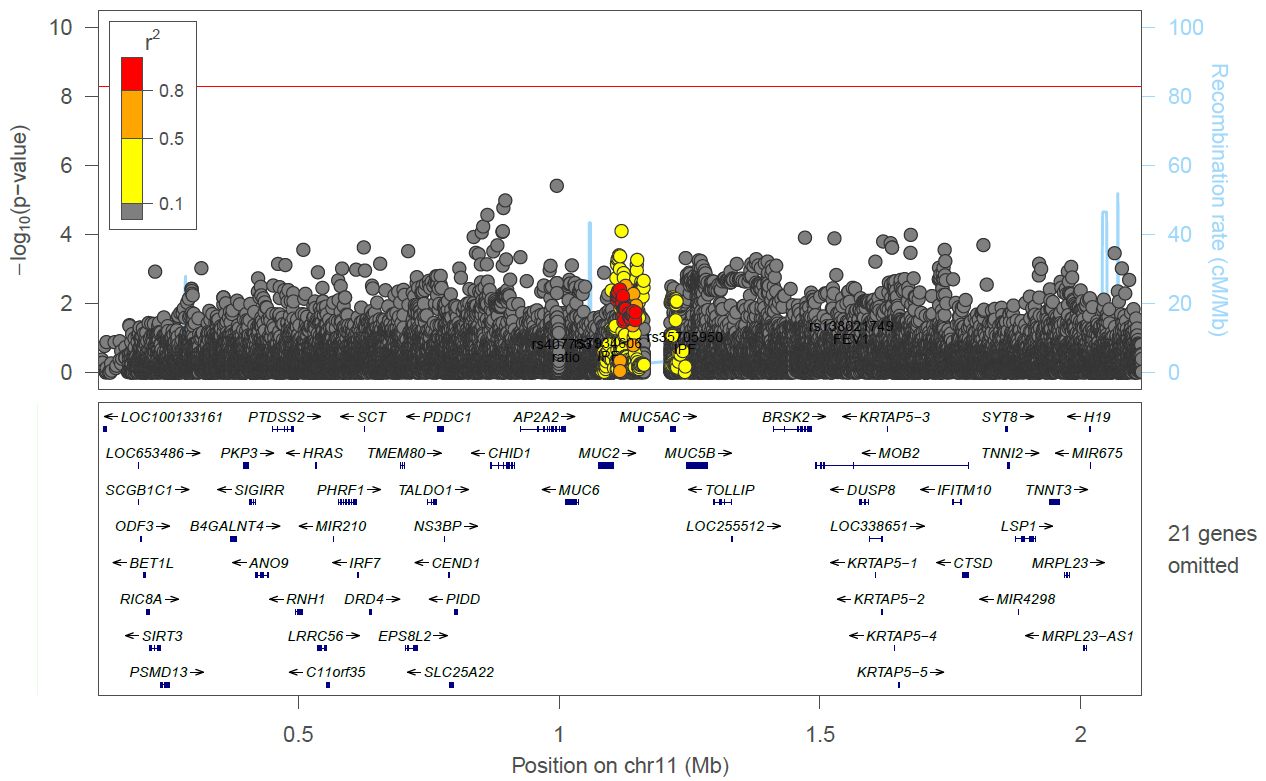

**Supplementary Figure 18:** Region plot of **rs779167905 signal** conditioned on moderate to severe asthma signal **rs11603634** (r^2^ = 0.451 with chronic sputum sentinel rs779167905, after conditioning chronic sputum P = 0.0039)

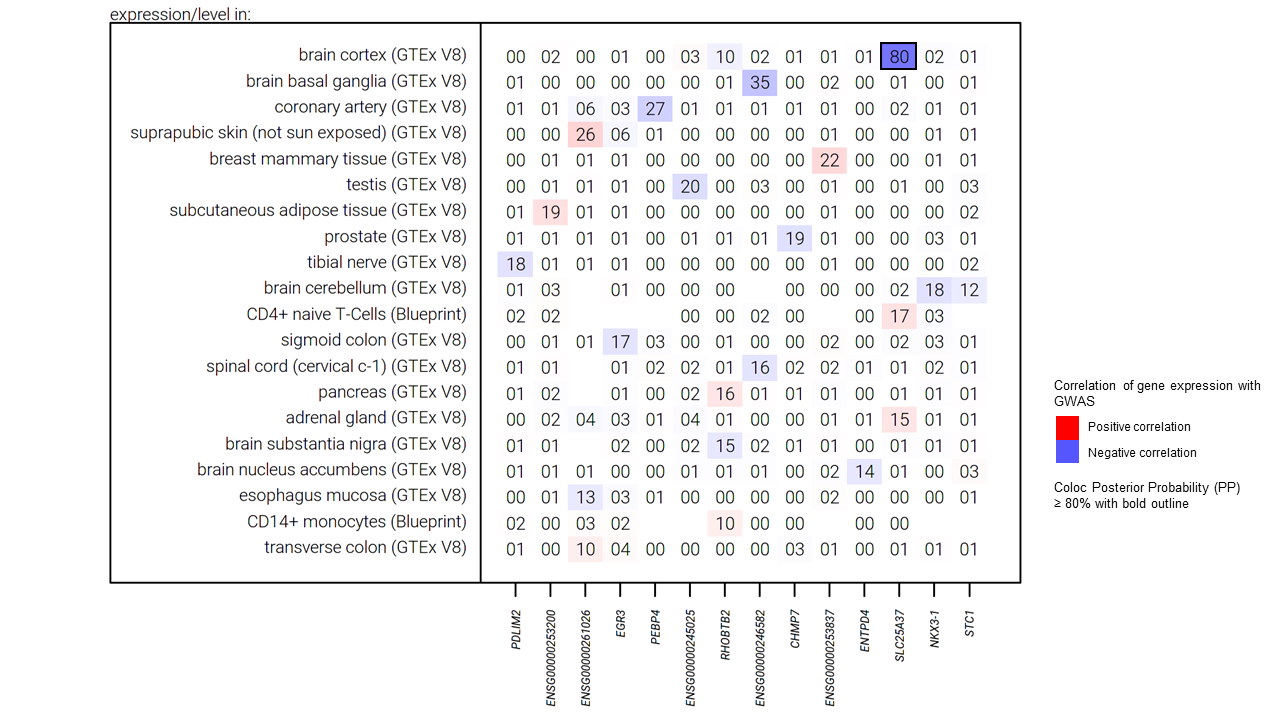

**Supplementary Figure 19:** Results for eQTL colocalization for the *NKX3*-1 locus using variant **rs79401075.** The numbers within the table are the posterior probability of colocalization (H4), with results aligned to the risk allele A for the **rs79401075** variant. Missing numbers indicate no data was available for the respective gene and tissue.

### Supplementary Table Legends

**Supplementary Table 1:** Diagnostic codes used for bronchiectasis. Source of codes V3=Read V3 codes, V2=Read V2 codes, ICD10=International classification of disease 10th edition, ICD9 = International classification of disease 10th edition, MD = Mortality data (ICD10 code), SR = Self report UK Biobank code (data-field 20002). Read V3 and Read V2 are primary care codes extracted from the primary care records. ICD10 codes are extracted from the hospital episodic statistics and mortality data.

**Supplementary Table 2:** Diagnostic codes used for cystic fibrosis. Source of codes V3=Read V3 codes, V2=Read V2 codes, ICD10=International classification of disease 10th edition, ICD9 = International classification of disease 10th edition, MD = Mortality data (ICD10 code). Read V3 and Read V2 are primary care codes extracted from the primary care records. ICD10 codes are extracted from the hospital episodic statistics and mortality data.

**Supplementary Table 3:** eQTL tissues and data source searched.

**Supplementary Table 4a:** Demographics, ever smoking status (UK Biobank data-field 22016), doctor diagnosed asthma (UK Biobank data-field 22127), doctor diagnosed chronic bronchitis (UK Biobank data-field 22129), cough and moderate to severe asthma status of those who answered yes or no to the question “do you bring up phlegm/sputum/mucus daily?” and were viable cases or controls.

**Supplementary Table 4b:** COPD status based on spirometry of cases and controls (spirometry-derived phenotypes are only available in those with spirometry data that passed quality control).

**Supplementary Table 4c:** Bronchiectasis and cystic fibrosis status of cases and controls (primary care, secondary care, mortality and self-reported phenotypes). Only individuals with linked primary care data included in the comparison.

**Supplementary Table 5:** Subgroup analysis for sentinel SNPs in sensitivity analyses. #CHROM = Chromosome, POS = position, REF = reference allele, ALT = alternative allele, CT = count, SE = standard error, A1 is the tested allele.

**Supplementary Table 6:** Results for GWAS look-ups, results aligned the chronic sputum production risk allele for all loci.

**Supplementary Table 7:** PheWAS results, all results aligned to risk allele for chronic sputum production. HLA-DRB1 results obtained from Deep-PheWAS. SE = standard error, FRD=false discovery rate, only results with FDR <0.01 are reported.

**Supplementary Table 8:** Associations with previously reported GWAS in Open Targets Genetics Portal. PMID=Pubmed ID. Only results with P<5x10^-8^ reported. OR=Odds ratio, P=p-value.

**Supplementary Table 9:** Ensemble variant effect predictor (VEP) results for *FUT2* locus.

**Supplementary Table 10:** Results for credible sets for each of the 5 non HLA signals, results aligned to chronic sputum production risk allele for all variants.

**Supplementary Table 11:** eQTL co-localisation results, results aligned to risk allele for chronic sputum production. H4 = posterior probability of the signals being the same.

**Supplementary Table 12:** Read V3 codes for sputum in original sample and stratified by current smoking status (field-ID 22506). Read V3 are primary care codes extracted from the primary care records. * =Overall N_cases = 4498, N_controls = 45403, **= Overall N_cases = 440, N_controls = 1509, ***= Overall N_cases = 4041, N_controls = 43676.

**Supplementary Table 13:** Results and meta-analysis from the five additional studies for the six sentinel variants. *= rs10902094 proxy, **= rs143032234 proxy, ª= rs504963 proxy. P=p-value, OR= Odds ratio, CI= Confidence Interval.
