## Supplementary material for "Genome-wide association study of chronic sputum production implicates loci involved in mucus production and infection": STEGA Strobe checklist

### TABLE 1. STREGA reporting recommendations, extended from STROBE Statement

| **Item** | **Item number** | **STROBE Guideline** | **Extension for Genetic Association Studies (STREGA)** | **Relevant text from manuscript** |
| --- | --- | --- | --- | --- |
| **Title and Abstract** | 1 | (a) Indicate the study’s design with a commonly used term in the title or the abstract. |  | Title and Abstract: “genome-wide association study” |
|  |  | (b) Provide in the abstract an informative and balanced summary of what was done and what was found. |  | Abstract: Methods (including sample size, significance threshold used) and Findings |
| **Introduction** | | |  |  |
| *Background rationale* | 2 | Explain the scientific background and rationale for the investigation being reported. |  | Abstract: Background  Introduction |
| *Objectives* | 3 | State specific objectives, including any pre-specified hypotheses. | ***State if the study is the first report of a genetic association, a replication effort, or both.*** | Introduction: Paragraph 4 – first report of genetic associations for chronic sputum production. |
| **Methods** | | |  |  |
| *Study design* | 4 | Present key elements of study design early in the paper. |  | Abstract: Methods and Findings. Hypothesis and overall approach outlined at end of Introduction.  Methods |
| *Setting* | 5 | Describe the setting, locations and relevant dates, including periods of recruitment, exposure, follow-up, and data collection. |  | Methods: Subjects  Data Availability |
| *Participants* | 6 | (a) **Cohort study –** Give the eligibility criteria, and the sources and methods of selection of participants. Describe methods of follow-up.   **Case-control study –** Give the eligibility criteria, and the sources and methods of case ascertainment and control selection. Give the rationale for the choice of cases and controls.  **Cross-sectional study –** Give the eligibility criteria, and the sources and methods of selection of participants. | ***Give information on the criteria and methods for selection of subsets of participants from a larger study, when relevant***. | Case-control study  Methods: Study population |
|  |  | (b) **Cohort study –** For matched studies, give matching criteria and number of exposed and unexposed.   **Case-control study –** For matched studies, give matching criteria and the number of controls per case. |  | Case-control study  Methods: Study population |
| *Variables* | 7 | *(a)* Clearly define all outcomes, exposures, predictors, potential confounders, and effect modifiers. Give diagnostic criteria, if applicable. | ***(b)*** ***Clearly define genetic exposures (genetic variants) using a widely-used nomenclature system. Identify variables likely to be associated with population stratification (confounding by ethnic origin).*** | Methods: Genome-wide association study of chronic sputum production  Rsids used to describe associated variants. |
| *Data sources measurement* | 8***** | *(a)* For each variable of interest, give sources of data and details of methods of assessment (measurement). Describe comparability of assessment methods if there is more than one group. | ***(b)*** ***Describe laboratory methods, including source and storage of DNA, genotyping methods and platforms (including the allele calling algorithm used, and its version), error rates and call rates. State the laboratory/centre where genotyping was done****.* ***Describe comparability of laboratory methods if there is more than one group. Specify whether genotypes were assigned using all of the data from the study simultaneously or in smaller batches.*** | Methods: Genome-wide association study of chronic sputum production |
| *Bias* | 9 | *(a)* Describe any efforts to address potential sources of bias. | ***(b) For quantitative outcome variables, specify if any investigation of potential bias resulting from pharmacotherapy was undertaken. If relevant, describe the nature and magnitude of the potential bias, and explain what approach was used to deal with this.*** | Sensitivity analyses undertaken to investigate contribution of pre-existing respiratory disease or smoking status, time of year for completion of survey, sex, current smoking vs ever smoking. |
| *Study size* | 10 | Explain how the study size was arrived at. |  | Methods: Study population |
| *Quantitative variables* | 11 | Explain how quantitative variables were handled in the analyses. If applicable, describe which groupings were chosen, and why. | ***If applicable, describe how effects of treatment were dealt with.*** | Methods: Genome-wide association study of chronic sputum production |
| Statistical methods | 12 | (a) Describe all statistical methods, including those used to control for confounding. | ***State software version used and options (or settings) chosen.*** | Methods: Genome-wide association study of chronic sputum production |
|  |  | (b) Describe any methods used to examine subgroups and interactions. |  | Methods: Associations with other phenotypes |
|  |  | (c) Explain how missing data were addressed. |  | Methods: Study population |
|  |  | (d) **Cohort study –** If applicable, explain how loss to follow-up was addressed.   **Case-control study –** If applicable, explain how matching of cases and controls was addressed.  **Cross-sectional study –** If applicable, describe analytical methods taking account of sampling strategy. |  | Methods: Study population |
|  |  | (e) Describe any sensitivity analyses. |  | Methods: Associations with other phenotypes |
|  |  |  | ***(f) State whether Hardy-Weinberg equilibrium was considered and, if so, how****.* | Methods: Genome-wide association study of chronic sputum production (Paragraph 1) |
|  |  |  | ***(g) Describe any methods used for inferring genotypes or haplotypes.*** | Methods: Genome-wide association study of chronic sputum production (Paragraph 1) |
|  |  |  | ***(h) Describe any methods used to assess or address population stratification.*** | Methods: Genome-wide association study of chronic sputum production |
|  |  |  | ***(i) Describe any methods used to address multiple comparisons or to control risk of false positive findings.*** | Methods: Genome-wide association study of chronic sputum production |
|  |  |  | ***(j) Describe any methods used to address and correct for relatedness among subjects*** | Methods: Study population |
| **Results** | | |  |  |
| *Participants* | 13***** | (a) Report the numbers of individuals at each stage of the study – e.g., numbers potentially eligible, examined for eligibility, confirmed eligible, included in the study, completing follow-up, and analysed. | ***Report numbers of individuals in whom genotyping was attempted and numbers of individuals in whom genotyping was successful.*** | Results: Paragraph 1, Table 1, figure 1 |
|  |  | (b) Give reasons for non-participation at each stage. |  | Results: Paragraph 1, figure 1 |
|  |  | (c) Consider use of a flow diagram. |  | Described in: Methods: Study population, figure 1 |
| *Descriptive data* | 14***** | (a) Give characteristics of study participants (e.g., demographic, clinical, social) and information on exposures and potential confounders. | ***Consider giving information by genotype****.* | Results: Table 1 |
|  |  | (b) Indicate the number of participants with missing data for each variable of interest. |  | Results: Table 1 |
|  |  | (c) **Cohort study –** Summarize follow-up time, e.g. average and total amount. |  | Not Applicable |
| *Outcome data* | 15 ***** | **Cohort study-**Report numbers of outcome events or summary measures over time. | ***Report outcomes (phenotypes) for each genotype category over time*** |  |
|  |  | **Case-control study –** Report numbers in each exposure category, or summary measures of exposure. | ***Report numbers in each genotype category*** | Not applicable for genome-wide study. Coded allele frequencies for variants of interest reported throughout. |
|  |  | **Cross-sectional study –** Report numbers of outcome events or summary measures. | ***Report outcomes (phenotypes) for each genotype category*** |  |
| *Main results* | 16 | (a) Give unadjusted estimates and, if applicable, confounder-adjusted estimates and their precision (e.g., 95% confidence intervals). Make clear which confounders were adjusted for and why they were included. |  | Results: Table 2 |
|  |  | (b) Report category boundaries when continuous variables were categorized. |  | Not applicable |
|  |  | (c) If relevant, consider translating estimates of relative risk into absolute risk for a meaningful time period. |  | Not applicable |
|  |  |  | ***(d) Report results of any adjustments for multiple comparisons.*** | Results: Table 2, Figure 4. |
| *Other analyses* | 17 | (a) Report other analyses done – e.g., analyses of subgroups and interactions, and sensitivity analyses. |  | Results: page 7, paragraph 1, Supplementary Figures 7 and 8, Supplementary Table 5. |
|  |  |  | ***(b) If numerous genetic exposures (genetic variants) were examined, summarize results from all analyses undertaken.*** | Results: Figure 2 |
|  |  |  | ***(c) If detailed results are available elsewhere, state how they can be accessed.*** | Data Availability |
| **Discussion** | | |  |  |
| *Key results* | 18 | Summarize key results with reference to study objectives. |  | Discussion: Paragraph 1 |
| *Limitations* | 19 | Discuss limitations of the study, taking into account sources of potential bias or imprecision. Discuss both direction and magnitude of any potential bias. |  | Discussion:  Paragraph 7 |
| *Interpretation* | 20 | Give a cautious overall interpretation of results considering objectives, limitations, multiplicity of analyses, results from similar studies, and other relevant evidence. |  | Discussion:  Paragraph 8 |
| *Generalizability* | 21 | Discuss the generalizability (external validity) of the study results. |  | Discussion:  Paragraph 6 |
| **Other Information** | | |  |  |
| *Funding* | 22 | Give the source of funding and the role of the funders for the present study and, if applicable, for the original study on which the present article is based. |  | Acknowledgements |

STREGA = STrengthening the REporting of Genetic Association studies; STROBE = STrengthening the Reporting of Observational Studies in Epidemiology.

* Give information separately for cases and controls in case-control studies and, if applicable, for exposed and unexposed groups in cohort and cross-sectional studies.
